# Quantification of Optical Coherence Tomography Features in >3500 Patients with Inherited Retinal Disease Reveals Novel Genotype-Phenotype Associations

**DOI:** 10.1101/2025.07.03.25330767

**Authors:** William Woof, Thales A. C. de Guimarães, Saoud Al-Khuzaei, Malena Daich Varela, Mital Shah, Gunjan Naik, Sagnik Sen, Pallavi Bagga, Bernardo Mendes, Yiu Wai Chan, Siying Lin, Biraja Ghoshal, Bart Liefers, Dun Jack Fu, Michalis Georgiou, Alan Sousa da Silva, Quang Nguyen, Yichen Liu, Dayyanah Sumodhee, Yu Fujinami-Yokokawa, Praveen J. Patel, Jennifer Furman, Ismail Moghul, Mariya Moosajee, Juliana Sallum, Samantha R. De Silva, Birgit Lorenz, Philipp Herrmann, Frank G. Holz, Kaoru Fujinami, Andrew R Webster, Omar A. Mahroo, Susan M. Downes, Savita Madhusuhan, Konstantinos Balaskas, Michel Michaelides, Nikolas Pontikos

## Abstract

**Purpose:** To quantify spectral-domain optical coherence tomography (SD-OCT) images cross-sectionally and longitudinally in a large cohort of molecularly characterized patients with inherited retinal disease (IRDs) from the UK.

**Design:** Retrospective study of imaging data.

**Participants:** Patients with a clinical and molecularly confirmed diagnosis of IRD who have undergone macular SD-OCT imaging at Moorfields Eye Hospital (MEH) between 2011 and 2019. We retrospectively identified 4,240 IRD patients from the MEH database (198 distinct IRD genes), including 69,664 SD-OCT macular volumes.

**Methods:** Eight features of interest were defined: retina, fovea, intraretinal cystic spaces (ICS), subretinal fluid (SRF), subretinal hyper-reflective material (SHRM), pigment epithelium detachment (PED), ellipsoid zone loss (EZ-loss) and retinal pigment epithelium loss (RPE-loss). Manual annotations of five b-scans per SD-OCT volume was performed for the retinal features by four graders based on a defined grading protocol. A total of 1,749 b-scans from 360 SD-OCT volumes across 275 patients were annotated for the eight retinal features for training and testing of a neural-network-based segmentation model, AIRDetect-OCT, which was then applied to the entire imaging dataset.

**Main Outcome Measures:** Performance of AIRDetect-OCT, comparing to inter-grader agreement was evaluated using Dice score on a held-out dataset. Feature prevalence, volume and area were analysed cross-sectionally and longitudinally.

**Results:** The inter-grader Dice score for manual segmentation was ≥90% for retina, ICS, SRF, SHRM and PED, >77% for both EZ-loss and RPE-loss. Model-grader agreement was >80% for segmentation of retina, ICS, SRF, SHRM, and PED, and >68% for both EZ-loss and RPE-loss. Automatic segmentation was applied to 272,168 b-scans across 7,405 SD-OCT volumes from 3,534 patients encompassing 176 unique genes. Accounting for age, male patients exhibited significantly more EZ-loss (19.6mm^2^ vs 17.9mm^2^, p<2.8×10^-4^) and RPE-loss (7.79mm^2^ vs 6.15mm^2^, p<3.2×10^-6^) than females. RPE-loss was significantly higher in Asian patients than other ethnicities (9.37mm^2^ vs 7.29mm^2^, p<0.03). ICS average total volume was largest in *RS1* (0.47mm^3^) and *NR2E3* (0.25mm^3^), SRF in *BEST1* (0.21mm^3^) and PED in *EFEMP1* (0.34mm^3^). *BEST1* and *PROM1* showed significantly different patterns of EZ-loss (p<10^-4^) and RPE-loss (p<0.02) comparing the dominant to the recessive forms. Sectoral analysis revealed significantly increased EZ-loss in the inferior quadrant compared to superior quadrant for *RHO* (Δ=-0.414 mm^2^, p=0.036) and EYS (Δ=-0.908 mm^2^, p=1.5×10^-4^). In *ABCA4* retinopathy, more severe genotypes (group A) were associated with faster progression of EZ-loss (2.80±0.62 mm^2^/yr), whilst the p.(Gly1961Glu) variant (group D) was associated with slower progression (0.56 ±0.18 mm^2^/yr). There were also sex differences within groups with males in group A experiencing significantly faster rates of progression of RPE-loss (2.48 ±1.40 mm^2^/yr vs 0.87 ±0.62 mm^2^/yr, p=0.047), but lower rates in groups B, C, and D.

**Conclusions:** AIRDetect-OCT, a novel deep learning algorithm, enables large-scale OCT feature quantification in IRD patients uncovering cross-sectional and longitudinal phenotype correlations with demographic and genotypic parameters.

## Introduction

Inherited Retinal Diseases (IRDs) are genetically and clinically heterogeneous monogenic disorders that affect the retina and represent the leading cause of legal blindness among working-age adults in England and Wales^1^, and the second commonest cause in childhood^2^.

The vast majority of IRDs are associated with structural changes within the retina, which can be detected with high-resolution retinal imaging using different imaging modalities. Spectral-domain (and swept source) Optical Coherence Tomography (SD-OCT) has become the most commonly used retinal imaging modality in ophthalmology and provides a highly sensitive non-invasive representation of *in vivo* retinal structure with strong correlation between SD-OCT and histopathology ^3,4^. Changes in optical reflectivity in SD-OCT imaging can demonstrate abnormalities of the vitreoretinal interface, neurosensory retina, retinal pigment epithelium (RPE), and choroid, and some of these patterns are highly characteristic of specific IRDs ^5–11^.

The identification and quantification of retinal disease-associated features within SD-OCT imaging can be critical for diagnosis, monitoring disease progression, providing prognostic information and assessing treatments in IRDs. The first step in quantifying retinal imaging-based biomarkers of disease involves identification and segmentation of these features. Manual segmentation is time-consuming and requires expert annotators, which makes this process subjective and infeasible on a large scale, especially in dense SD-OCT volume scans. Automated identification and segmentation of IRD features in a reliable way is important for enabling the routine use of these data quantitatively in clinical practice and research. Deep learning provides an effective way to automatically identify and segment features directly from imaging data. For example, automated methods for segmenting the ellipsoid zone (EZ) are of particular interest as this imaging feature represents regions of preserved photoreceptors, an important biomarker in clinical trials ^12,13^. Existing studies using deep learning for the automated identification and segmentation of SD-OCT features have so far been limited to specific IRD phenotypes or diagnoses^12,14–19^.

We recently published an AI algorithm (AIRDetect) that segments IRD features in fundus autofluorescence (FAF) imaging ^20^. To complement this approach, we present AIRDetect-OCT, an AI algorithm that can automatically identify and segment relevant features from SD-OCT images. The benefit of SD-OCT image analysis is that it provides a more detailed and comprehensive direct view of the 3D structure of the retina compared to 2D FAF images^20,21^. Additionally, manual annotation of SD-OCT volumes is substantially more laborious than FAF images, making automatic annotation vital even for smaller datasets. We present our results from applying AIRDetect-OCT to a large and heterogeneous cohort of over 3500 patients with molecularly confirmed IRDs from a single center to determine its performance, identify cross-sectional associations with demographic and genetic parameters, as well as quantify disease progression.

## Method

### Dataset curation

A similar process to our FAF study was followed^20^. Patients’ genotypes were extracted from the genetics database of Moorfields Eye Hospital (MEH, London, UK) ^22,23^. Patients’ images captured between 2011-03-04 and 2019-10-22 were exported from the Heidelberg Imaging database (Heidelberg Engineering, Heidelberg, Germany) based on their hospital number.

The total MEH IRD SD-OCT dataset consisted of 4,240 patients covering 198 genes corresponding to 69,664 SD-OCT volumes. From this, a dataset of 1749 SD-OCT b-scans corresponding to 360 SD-OCT volumes from 275 patients were manually annotated by four graders (TACG, MDV, SS, SAK) for eight different image features to be used for training and internal testing of the AI model as defined in **Table 1**. A grading protocol was defined to identify these annotations, which was refined iteratively (**Table 1)** ^24^. This data set was further split into train (237 patients, 1,389 b-scans) and test datasets (38 patients, 360 b-scans). A dataset of 3,534 patients, 176 genes, 7,405 SD-OCT volumes, and 272,168 individual b-scans was kept aside for analysis from the initial total dataset. The completed data flow is summarized in **Figure S1**.

**Table 1:** Description of the eight annotated retinal OCT features along with the instructions for manual annotation.

| Feature | Description | Instructions |
| --- | --- | --- |
| Retina | Full extent of the retina from inner limiting membrane (ILM) to the Bruch's membrane (BM) | The entire retina is annotated in a single zone which spans from BM to the ILM. |
| Intraretinal Cystic Spaces (ICS) | Dark circular or ovoid spaces located within the neuroretina and clearly distinguishable from the surrounding retinal tissue (alternatively known as intraretinal cystic spaces). Also includes schisis. | The intraretinal cystic spaces are annotated in their entirety, respecting the regions of integrity between each space. Intraretinal fluid that is diffusely accumulated within the neuroretina causing retinal thickening without clearly distinguishable cystic spaces should not be annotated. |
| Subretinal Fluid (SRF) | Subretinal fluid is defined as a homogenous hyporeflective space located in the subretinal space (between the neuroretina and the RPE). The outer boundary of the neuroretina is defined as the ellipsoid zone (EZ). | Subretinal fluid is to be annotated in its entirety. Any hyporeflective area in the subretinal space is to be annotated (not to confuse with optical gap caused by a local subfoveal disruption in the ellipsoid zone). In the presence of adjacent hyperreflective material, this is to be considered subretinal hyperreflective (SHRM) material and thus not to be annotated. |
| Subretinal Hyper-Reflective Material (SHRM) | Hyper-reflective material below the neuroretina and above the RPE. Includes vitelliform material in some genotypes, such as *BEST1*. | SHRM is to be annotated as any hyperreflective material present in the subretinal space. This requires accuracy as SHRM may overlap with areas of ellipsoid zone loss (EZ-loss) and SRF. |
| Pigment Epithelium Detachment (PED) | Separation of RPE from Bruch's membrane. These can be drusenoid, serous, or fibrovascular. | PED is to be annotated in its full extent. It may be difficult to visualize and delineate the exact borders of the PED, in which case the best estimation of the Bruch's membrane needs to be taken. |
| Ellipsoid Zone loss (EZ-loss) | Disruption/loss of EZ is known to be diagnostic and clinically significant. EZ-loss includes EZ+IZ loss (when IZ is discernible). The Interdigitation Zone (IZ) is another hyper-reflective layer discernible between EZ and RPE. | Similar to RPE-loss, this feature should be drawn in the relative area where the EZ would be expected to be. Only the horizontal coordinates are used to annotate this feature since the vertical coordinate (EZ thickness) is challenging to annotate due to the layer being normally thin and poorly reproducible. |
| Retinal Pigment Epithelium loss (RPE-loss) | Retinal Pigment Epithelium loss is known to be diagnostic and clinically significant. | Retinal Pigment Epithelium loss is annotated using only the horizontal coordinates of the feature where the RPE would be expected to be but is missing. In well-defined regions of RPE atrophy, such as cases of *ABCA4*-associated retinal dystrophy, a hypertransmission zone may be present in the choroid, which can guide the grading. |
| Foveal Centre | Foveal pit | The foveal pit may span a number of b-scans, but only one b-scan should be annotated with the fovea center. A useful clue for determining the foveal center is the appearance of a brighter Internal Limiting Membrane (ILM) in one b-scan only - this is the b-scan where the foveal center is located. |

All OCT volumes were macular centered non-radial images captured by the Heidelberg Spectralis imaging platform. The Dice similarity coefficient score across the pairs of individually segmented masks was used to assess inter-grader agreement ^25^, except for retinal pigment epithelium loss (RPE-loss) and ellipsoid zone loss (EZ-loss) where only the horizontal overlap was taken into account. To enable a greater number of SD-OCT volumes to be annotated only five b-scans per SD-OCT volume were annotated: the first and last b-scan in the volume, the foveal b-scan, as well as the two adjacent b-scans two steps directly above and below the foveal b-scan. This approach provided greater data diversity compared to annotating all b-scans for a smaller number of volumes.

Manual grading was completed over an 18-month period from June 2022 to December 2023 by four graders, concurrently with grading for AIRDetect FAF^20^. Grading was performed using the Moorfields Grading Portal online platform (grading.readingcentre.org) by research fellows with more than five years’ experience in medical retina and three with IRD expertise. In total, after discarding ungradable and incomplete gradings, 1,749 b-scans were graded across 275 patients and 75 unique genes, of which 503 were double-graded (81 patients, 30 unique genes).

### Model Training and Evaluation

Four separate segmentation models were trained and combined into one AIRDetect-OCT model. One for Retina, one for PED/SHRM/SRF/ICS features, one for RPE/EZ-loss, and one for Fovea. Prior to training we selected a held-out test set of 38 patients (across 31 unique genes), consisting of 360 annotated b-scans, which were excluded from all training and development of all models. For training and evaluation, for double graded scans annotations were selected randomly from available annotations.

For the Retina segmentation model, the U-net model with X-ception backbone was selected, using the general Imagenet weights as pretrained weights for the encoder^26^. As the Retina feature is present in all available SD-OCT b-scans, a standard binary cross entropy loss function along with Adam optimizer was used for training. The model was trained for 200 epochs with horizontal flipping, histogram equalization and 10-degree rotations for image augmentation.

For the PED/SHRM/SRF/ICS segmentation model, the nnU-Net (no-new-UNet) framework was selected for its adaptability and performance in automatic medical image segmentation tasks ^27^. A single multi-class model for all four features was trained using sum of dice and cross-entropy loss functions to optimize multi-class segmentation accuracy. Hyperparameters, such as learning rate and batch size, were selected by the nnU-Net based on its analysis of the dataset. Training was curtailed at 1000 epochs as this was found to be sufficient to achieve convergence. Cross-validation was applied to prevent overfitting and obtain a more generalizable model.

For the RPE/EZ-loss model, a fine-tuning approach was applied. The pretrained weights from a geographic atrophy segmentation study were used for defining EZ-loss and RPE-loss which uses a Unet with a Resnet backbone ^28^. Data augmentation included horizontal flipping along with rotations of ± 10 degrees. Dice loss along with Adam optimizer were used for training. The model was trained for 200 epochs considering the smaller dataset size and use of pretrained weights. A minimum area threshold was set for these features using a heuristic approach with 120 pixels for EZ-loss and 300 for RPE-loss.

For the Fovea segmentation model, a U-net with Resnet architecture with focal loss was used as the fovea is annotated as a single point on an SD-OCT b-scan, therefore the class imbalance between background pixels and fovea pixel is high. The training data consisted of b-scans on which the fovea was present annotated with their x,y location (in pixels) on the relevant b-scan. To improve training efficiency and reduce data imbalance, one b-scan per SD-OCT volume was randomly selected as opposed to including all b-scans per volume. The model was trained for 2500 epochs. During inference, the probability maps for all b-scans in a given volume were calculated and the highest probability value was selected from the resulting 3D array.

Model performance was assessed using the Dice coefficient between the model predictions and the corresponding grader annotation on the hold-out test set. For EZ-loss and RPE-loss we considered only the horizontal extent when calculating Dice, using the 1-dimensional dice of the flattened representation of both masks. Where b-scans were double graded, we took the mean of the model-grader Dice for each grading. We also analyzed the model-grader agreement for simple whole-image presence/absence, where we counted cases as positive for which the annotator marked at least some part of the image for the given feature, and negative otherwise, from which we derived accuracy, precision and recall. We also assessed inter-grader agreement by using the dice between annotations of double-graded scans.

### Applying automatic annotations to the MEH cohort

All models were applied to the full MEH dataset consisting of 69,664 OCT volumes from 4,240 patients across 198 unique IRD genes.

To ensure greater consistency between volumes, volumes with enface dimensions smaller than 4.5mm, or larger than 7.5mm in either the height or the width dimension were excluded. Median volume enface dimensions were 5.86mm x 5.86mm. After filtering, 1,521,757 images from 28,348 SD-OCT volumes from 3534 patients across 176 genes remained. For cross-sectional analysis, only SD-OCT volumes from patients at first presentation were included for analysis.

The number of b-scans per volume ranged between 6 and 193, with median and mode of 25 (**Table S3**). After filtering, 272,168 images from 7,405 SD-OCT volumes from 3,534 patients across 176 different IRD genes remained from the cross-sectional dataset (**Figure S1**).

For each of the generated masks we extracted: a) the area, number of pixels in the segmented mask multiplied by the resolution; b) the horizontal extent, the total span of the feature across the width of the b-scan. These were used to derive the total volume and enface area of each SD-OCT volume, by aggregating these values across b-scans and multiplying by the per-b-scan spacing. Features were also analyzed based on their distance from the fovea. Calculations were performed using the PyeScan python library (https://github.com/pontikos-lab/PyeScan/).

Rate of progression was calculated per patient-eye, where a linear regression was fit to the data, with the independent variable being the scan date since baseline (in years) and the dependent variable being the target feature (e.g. total area of EZ-loss). The resulting slope of the regression line fit to all timepoints per patient-eye was taken as the rate of progression. These per-eye rates were then averaged across the relevant cohort to get a per-group mean. To ensure greater consistency, only patients with imaging covering an interval of at least 365 days were included. Additionally, for EZ-loss and RPE-loss only patients with a baseline (initial) value of between 5 and 25 mm^2^ were included to avoid floor and ceiling effects.

## Results

### AIRDetect-OCT Model Validation

Examples of AIRDetect-OCT segmentation output are presented in **Figure 1**. Inter-grader agreement for manual segmentation was very high for Retina, ICS, SRF, SHRM and PED with inter-grader Dice scores greater than or equal to 90% (**Table 2**), though in part this was due to low feature incidences. Inter-grader agreement for manual segmentation of EZ-loss was 84.7%, and for RPE-loss was 77.4% (**Table 2**).

**Figure 1:**
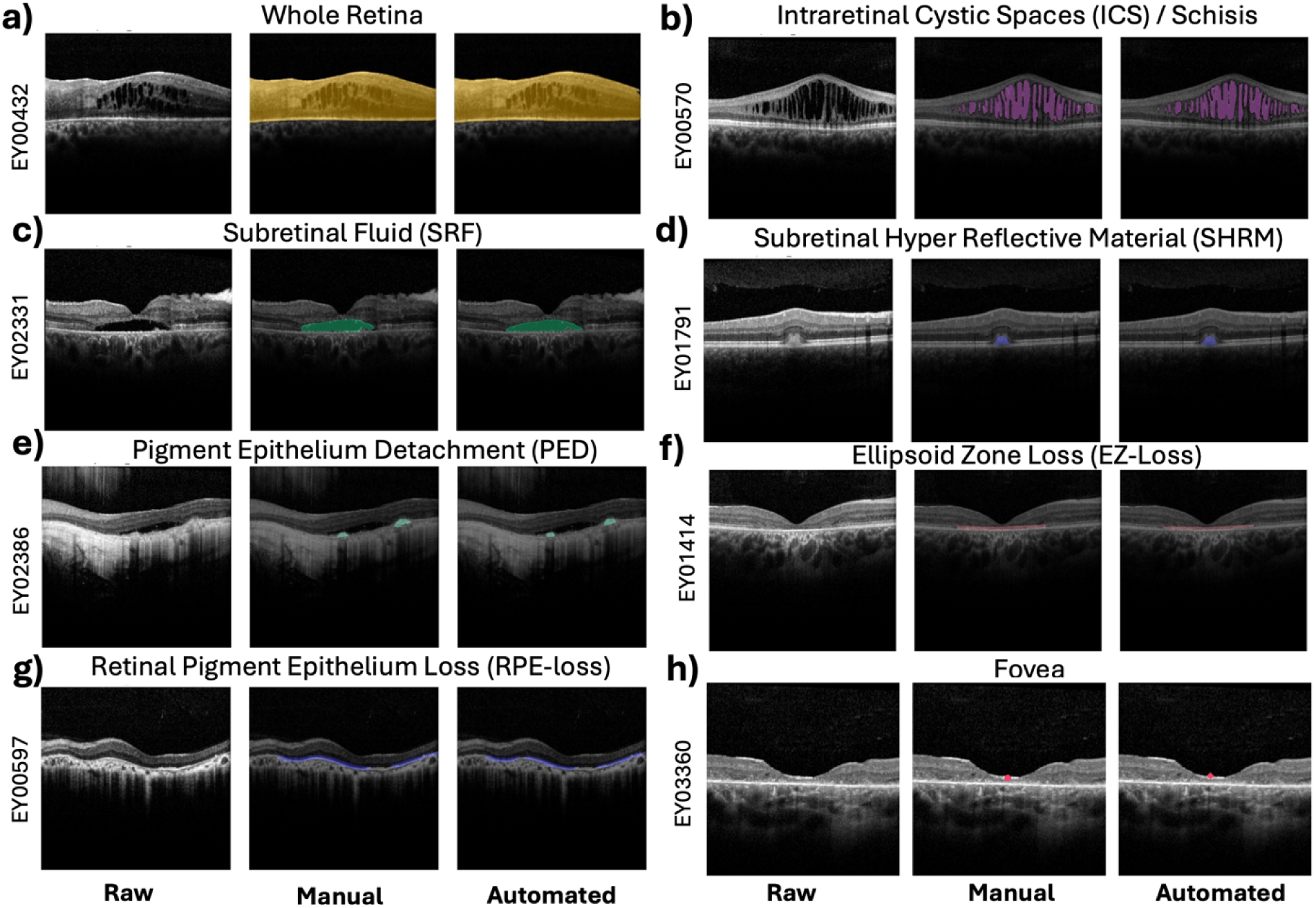
Example segmented masks of the eight features which include (**a**) whole retina, (**b**) intraretinal cystic spaces (ICS), (**c**) subretinal fluid (SRF), (**d**) subretinal hyper-reflective material (SHRM), (**e**) pigment epithelium detachment (PED), (**f**) ellipsoid zone loss (EZ-loss), (**g**) retinal pigment epithelium loss (RPE-loss) and (**h**) fovea localisation.

**Table 2:** Segmentation model training data and results. Dice score quantifies the model’s segmentation performance and presence/absence quantifies its feature detection performance. Total = number of annotated images. Incidence = percent of images with gradable features. Dice inter-grader = inter-grader agreement of double-graded images. Dice model-grader = agreement between model and graders, with mean scores used when images were double-graded.

| Feature | Train set |  | Test Set (n=360) | Segmentation (Dice) |  | Detection (Presence/Absence) |  |  |
| --- | --- | --- | --- | --- | --- | --- | --- | --- |
|  | Total | Incidence | Incidence | Inter-grader | Model-grader | Accuracy | Precision | Recall |
| Retina | 627* | - | - | 98.0% | 95.0% | - | - | - |
| ICS | 949 | 25.1% | 15.6% | 96.2% | 89.2% | 93.6% | 76.2% | 85.7% |
| SRF | 949 | 10.9% | 11.9% | 97.8% | 92.3% | 95.8% | 79.2% | 88.4% |
| PED | 949 | 16.7% | 10.6% | 90.0% | 81.1% | 84.2% | 35.8% | 63.2% |
| SHRM | 949 | 13.7% | 14.2% | 92.0% | 85.7% | 92.2% | 72.5% | 72.5% |
| EZ-loss | 1031 | 69.4% | 57.2% | 84.7%** | 69.2%** | 78.1% | 74.9% | 92.7% |
| RPE-loss | 1031 | 35.4% | 38.3% | 77.4%** | 68.7%** | 76.4% | 68.8% | 70.3% |

\* Retina model was trained before all grading was completed
\*\* Dice for EZ-loss and RPE-loss was calculated using horizontal extent only, ignoring vertical position.

There was a very high level of model-grader agreement (Dice>90%) for segmentation of Retina and SRF, and high model-grader agreement (Dice>80%) for ICS, SHRM, and PED (**Table 2**). There was a lower level of model-grader agreement for segmentation of RPE-loss (68.7%) and EZ-loss (69.2%) **(Table 2)**.

The model’s ability to correctly identify the presence or absence of features within images (accuracy) ranged between 76.4% to 95.8% with the lowest being RPE-loss, and highest being SRF (**Table 2**). Precision (also known as positive predictive value) measures the proportion of images where the feature was marked as present by the model which were also marked as present by the human grader and ranged from 68.8% - 79.2% except for PED which was notably lower, at 35.8% (**Table 2**). Recall (also known as sensitivity) measures the proportion of correctly identified features from the total number of these features in the dataset as annotated by the graders, and ranged from 70.3% to 92.7%, except PED which was 63.2% (**Table 2**).

### Demographic-phenotype correlations

Comparing across patient demographics we observe that male patients exhibit significantly higher EZ-(p<2.9×10^-4^), RPE-loss (p<3.3×10^-7^) and ICS (p<7.6×10^-5^) than female patients (**Table 3**). Asian and Asian British patients also exhibited significantly greater average RPE-loss (p<0.05) when compared to all other patients (**Table 4**).

**Table 3:** Age, average feature thickness/volume/enface area by patient sex with 95% confidence interval.

| Sex | Age (mean) | Retina (mm) | ICS (mm<sup>3</sup>) | SRF (mm<sup>3</sup>) | SHRM (mm<sup>3</sup>) | PED (mm<sup>3</sup>) | EZ-loss (mm<sup>2</sup>) | RPE-loss (mm<sup>2</sup>) |
| --- | --- | --- | --- | --- | --- | --- | --- | --- |
| Female | 47.6 | 0.269<br>±0.002 | 0.0166<br>±0.0052 | 0.00732<br>±0.00304 | 0.00338<br>±0.00092 | 0.00838<br>±0.00294 | 17.9<br>±0.7 | 6.14<br>±0.45 |
| Male | 45.2 | 0.265<br>±0.002 | 0.0391<br>±0.0010 | 0.01240<br>±0.00436 | 0.00293<br>±0.00074 | 0.00608<br>±0.00175 | 19.6<br>±0.6 | 7.79<br>±0.44 |
| p-value | $1.7 \times 10^{-4}$ | $6.3 \times 10^{-3}$ | $7.5 \times 10^{-5}$ | 0.061 | 0.46 | 0.19 | $2.8 \times 10^{-4}$ | $3.2 \times 10^{-7}$ |

**Table 4:** Average feature thickness/volume/enface area by patient ethnicity with 95% confidence interval. P-value given by on-way ANOVA.

| Ethnicity | Age (mean) | Retina (mm) | ICS (mm<sup>3</sup>) | SRF (mm<sup>3</sup>) | SHRM (mm<sup>3</sup>) | PED (mm<sup>3</sup>) | EZ-loss (mm<sup>2</sup>) | RPE-loss (mm<sup>2</sup>) |
| --- | --- | --- | --- | --- | --- | --- | --- | --- |
| Asian or Asian British | 44.3 | 0.254<br>±0.005 | 0.0177<br>±0.0155 | 0.00978<br>±0.0088 | 0.00125<br>±0.00133 | 0.00331<br>±0.00186 | 20.9<br>±1.5 | 9.37<br>±1.21 |
| Black or Black British | 45.3 | 0.260<br>±0.010 | 0.0384<br>±0.0427 | 0.0331<br>±0.0374 | 0.00066<br>±0.00059 | 0.00379<br>±0.00181 | 17.9<br>±2.7 | 7.01<br>±1.91 |
| Mixed | 37.7 | 0.264<br>±0.015 | 0.0184<br>±0.0275 | 0.0119<br>±0.0223 | 0.00806<br>±0.0146 | 0.00370<br>±0.00277 | 19.3<br>±5.2 | 7.82<br>±3.97 |
| Other Ethnic Groups | 44.8 | 0.267<br>±0.005 | 0.0239<br>±0.0168 | 0.0167<br>±0.0168 | 0.00356<br>±0.00226 | 0.00591<br>±0.00543 | 18.7<br>±1.6 | 6.75<br>±0.08 |
| White | 48.8 | 0.267<br>±0.003 | 0.0352<br>±0.0142 | 0.00777<br>±0.00391 | 0.00293<br>±0.00094 | 0.00573<br>±0.00160 | 19.4<br>±0.8 | 7.58<br>±0.60 |
| **p-value** | 8.9×10<sup>-6</sup> | **0.0012** | 0.72 | 0.12 | 0.10 | 0.73 | 0.22 | **0.029** |

We also saw several effects correlate with patient age. We observe retinal thickness decreasing monotonically across age groups, from 289±6µm in 0-18 year old patients, to 246±3µm in the 65+ cohort. Similarly, ICS significantly decreased from 69.5±28.2×10^-3^mm^3^ to 14.9±7.7×10^-3^mm^3^. Meanwhile volume of PEDs increased significantly with age from 1.42±1.20×10^-3^mm^3^ to 11.96±5.45×10^-3^mm^3^. EZ- and RPE-loss also increased significantly with age with EZ increasing from 9.9±1.6mm in the youngest group of patients to 21.9±0.9mm in the oldest group, and RPE increasing from 1.23±0.37mm to 12.12±0.89mm. SRF and SHRM showed no overall trend with the highest levels of SRF in the 18-30 range (12.7±7.6×10^-3^ mm^3^) and highest levels of SHRM in the 0-18 range (5.99±3.90×10^-3^ mm^3^) (**Table 5**).

**Table 5:** Average feature thickness/volume/enface area by patient age with 95% confidence interval. Beta and p-value are given for regression of patient age versus feature value before bucketing.

| Age range | Retina (mm) | ICS (mm<sup>3</sup>) | SRF (mm<sup>3</sup>) | SHRM (mm<sup>3</sup>) | PED (mm<sup>3</sup>) | EZ-loss (mm<sup>2</sup>) | RPE-loss (mm<sup>2</sup>) |
| --- | --- | --- | --- | --- | --- | --- | --- |
| 0-18 | 0.289<br>±0.006 | 0.0695<br>±0.0282 | 0.0093<br>±0.0067 | 0.00599<br>±0.00390 | 0.00142<br>±0.00120 | 9.8<br>±1.6 | 1.23<br>±0.37 |
| 19-30 | 0.282<br>±0.004 | 0.0527<br>±0.0293 | 0.0127<br>±0.0076 | 0.00376<br>±0.00198 | 0.00205<br>±0.00096 | 15.3<br>±1.2 | 3.23<br>±0.57 |
| 31-45 | 0.272<br>±0.003 | 0.0292<br>±0.0107 | 0.0107<br>±0.0065 | 0.00213<br>±0.00084 | 0.00422<br>±0.00173 | 19.1<br>±0.9 | 6.00<br>±0.57 |
| 45-65 | 0.264<br>±0.002 | 0.0197<br>±0.0062 | 0.0088<br>±0.0042 | 0.00305<br>±0.00086 | 0.00985<br>±0.00343 | 20.1<br>±0.7 | 7.90<br>±0.54 |
| 66+ | 0.246<br>±0.004 | 0.0149<br>±0.0077 | 0.0103<br>±0.0060 | 0.00333<br>±0.00136 | 0.01196<br>±0.00545 | 21.9<br>±0.9 | 12.12<br>±0.89 |
| β | -7.00×10<sup>-4</sup><br>±0.79×10<sup>-4</sup> | -8.19×10<sup>-4</sup><br>±3.22×10<sup>-4</sup> | -0.27×10<sup>-4</sup><br>±1.52×10<sup>-4</sup> | -1.09×10<sup>-5</sup><br>±3.17×10<sup>-5</sup> | 2.22×10<sup>-4</sup><br>±0.89×10<sup>-4</sup> | 0.149<br>±0.023 | 0.172<br>±0.016 |
| **p-value** | **<10<sup>-15</sup>** | **6.6×10<sup>-7</sup>** | 0.72 | 0.50 | **9.2×10<sup>-7</sup>** | **<10<sup>-15</sup>** | **<10<sup>-15</sup>** |

### Feature prevalence per gene

Comparing prevalence of features across the 30 most common genes in our dataset, the highest volumes of ICS were in *RS1* (0.427 mm^3^±0.138 CI_95_) and *NR2E3* (0.253 mm^3^±0.260) patients **(Figure 2b**). *BEST1* was associated with the highest volumes of SRF (0.207 mm^3^ ±0.055) and SHRM (0.0448 mm^3^±0.0108). *EFEMP1* was associated with the highest volumes of PED (0.343 mm^3^±0.122) and second highest volumes of SHRM (0.0393 mm^3^±0.0241) (**Figure 2d**). Average retinal thickness was highest in *CRB1* (0.334 mm ± 0.014), *NR2E3* (0.324mm ±0.023), *BEST1* (0.321mm ±0.006), *RS1* (0.310mm ±0.012), and *EFEMP1* (0.303mm ±0.011), whereas lower average retinal thickness was seen in *RP2* (0.239mm ±0.016) and *CERKL* (0.244mm ±0.017) (**Figure 2a)**.

**Figure 2:**
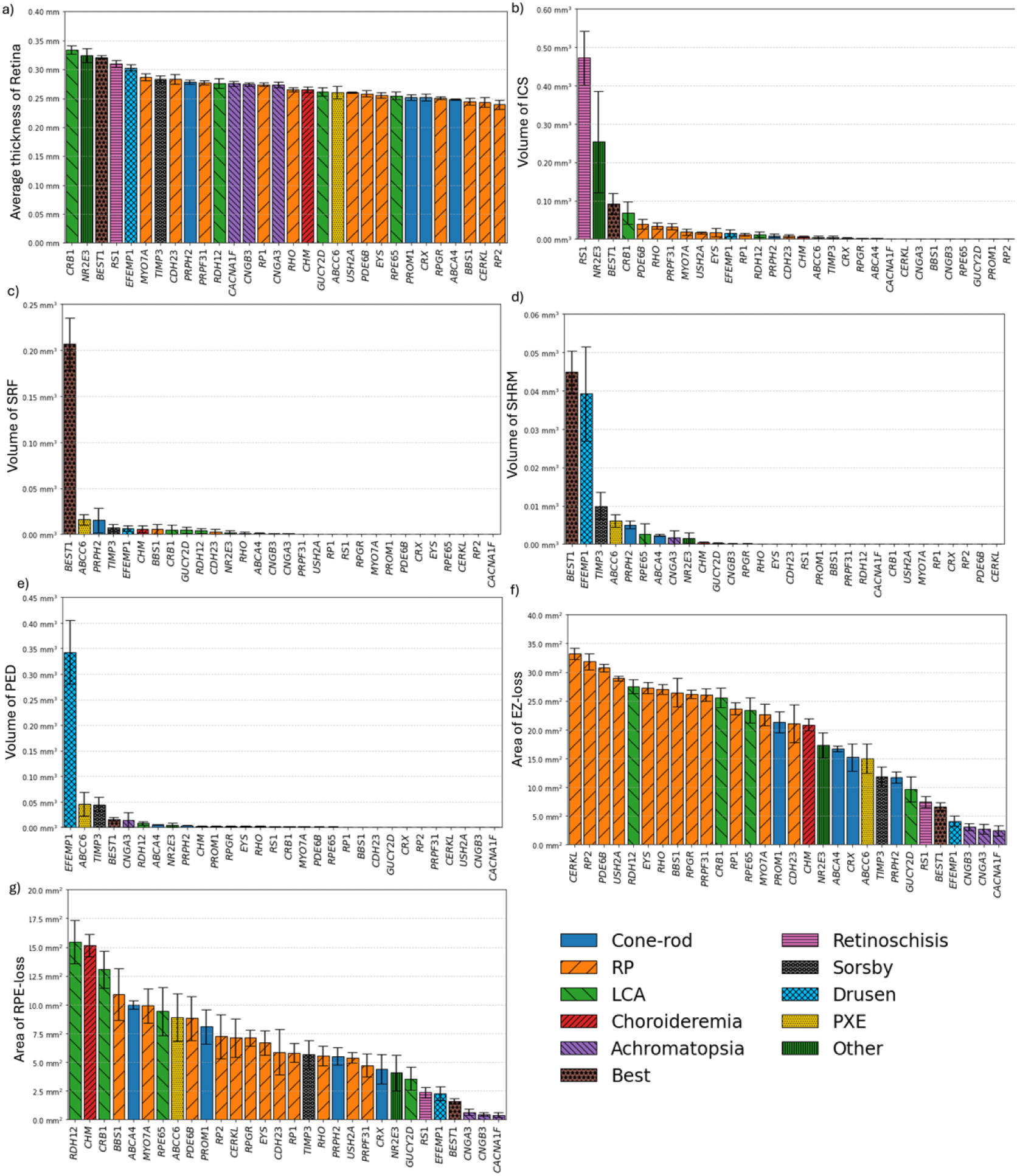
Average per patient (for initial appointment within our dataset) (**a**) Retinal thickness, volume of (**b**) ICS, (**c**) SRF, (**d**) SHRM, (**e**) PED/drusen, and enface area of (**f**) EZ-loss, (**g**) RPE-loss within SD-OCT volumes across 30 selected IRD genes. Error bars denote standard error (= std/sqrt(n)).

Across the 14 most common genes in our cohort associated with retinitis pigmentosa we found that *RPGR* (0.00196 mm^3^ ±0.00150) and *RP2* (0.0000351 mm^3^ ±0.0000347), were associated with very reduced volumes of ICS (**Figure 3b**). In genotypes associated with dominant inheritance patterns, except *PRPF8* which had high variability, ICS volumes were highest in *RHO* (0.0346 mm^3^ ±0.0166) and *PRPF31* (0.0324 mm^3^ ± 0.0166), and in recessively inherited genes ICS volumes were highest in *PDE6B* (0.0392 mm^3^ ± 0.0255) and *MYO7A* (0.0184 mm^3^ ±0.0162) (**Figure 3b**). Overall, we did not find a clear trend between recessive, dominant and X-linked RP genes in either of the four retinal features in **Figure 3**.

**Figure 3:**
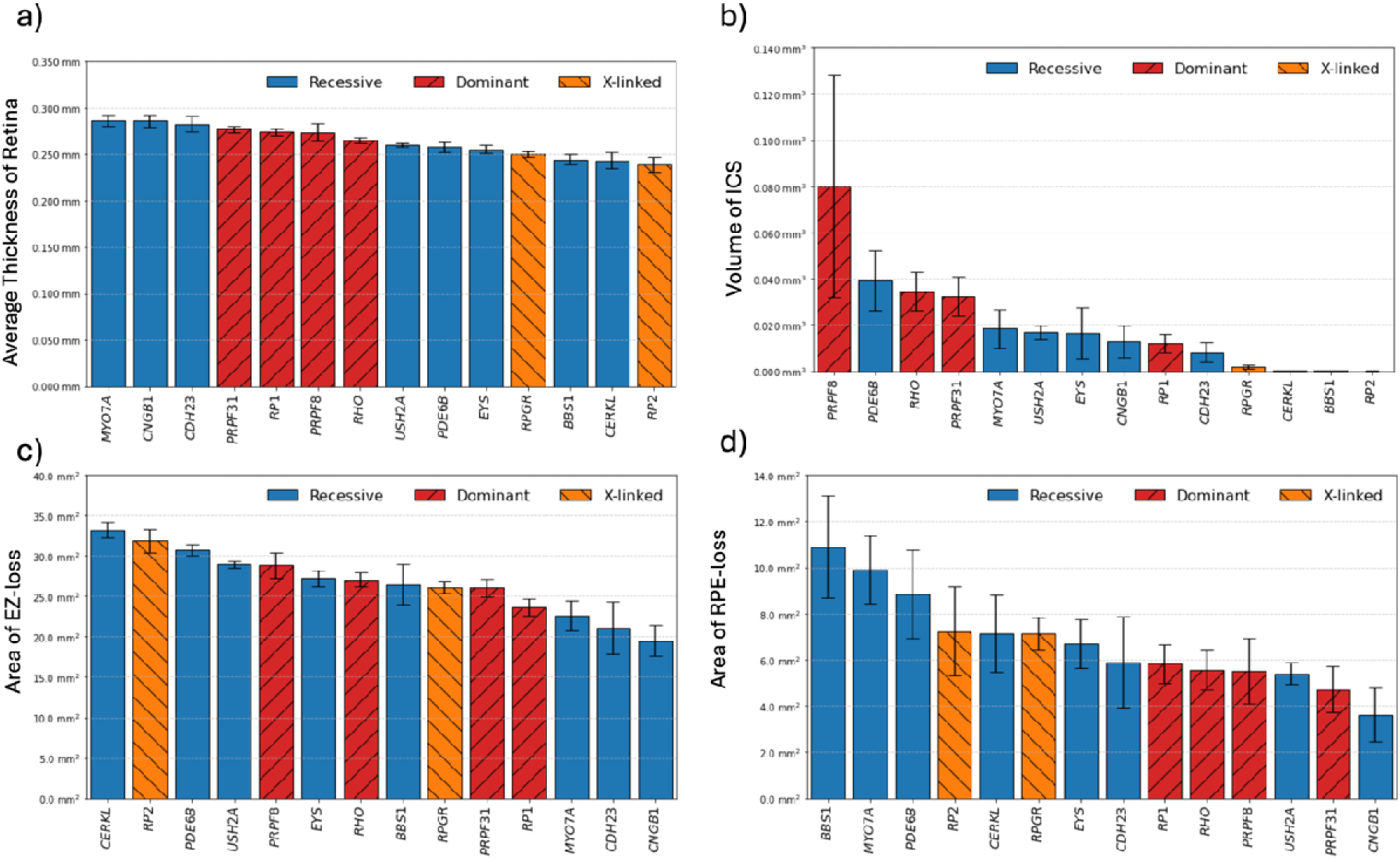
Average per patient (for initial appointment within our dataset) (**a**) Retinal thickness, volume of (**b**) ICS, and enface area of (**c**) EZ-loss (**d**) RPE-loss, within SD-OCT volume scans across genotypes associated with retinitis pigmentosa with at least 20 patients in our filtered dataset. Error bars denote standard error (= std/sqrt(n)).

Analyzing individual genes cross sectionally by age also gives some insight into progression patterns of different conditions (**Figure S2**). For example, EZ-loss appears to be less severe and exhibit later onset in *PRPH2* compared to *ABCA4*.

### Genotype-phenotype sectoral analysis

We compare quadrants (derived from en-face projection of OCT features) of the retina across genes to identify potentially novel phenotypic patterns (**Figure 5.b**). Across all patients, patients experienced significantly more EZ- and RPE-loss in the inferior quadrant than in the superior quadrant with a mean difference of Δ=0.056 mm^2^ (p=0.019) in EZ-loss, and Δ=0.099 mm^2^ (p=7.5×10^-9^) in RPE-loss.

Analyzing by genotype, *CNGA3, CNGB3* and *EYS* patients exhibited significantly more EZ-loss in the inferior quadrant than in the superior, with mean differences of Δ=-0.145 mm^2^ (p=0.024), Δ=-0.248 mm^2^ (p=0.0020), and Δ=-0.91 mm^2^ (p=1.5×10^-4^) respectively. *CHM* (Δ=0.327 mm^2^, p=0.019), and *PRPH2* (Δ=0.227 mm^2^, p=0.023) patients exhibited greater superior RPE-loss, while *EYS* (Δ=-0.502 mm^2^, p=2.0×10^-4^), *PDE6B* (Δ=-0.380 mm^2^, p=0.0031), and *RPGR* (Δ=0.334 mm^2^, p=5.3×10^-5^) patients exhibited greater inferior RPE-loss.

Laterally, numerous genes exhibited differences between nasal and temporal quadrants with *ABCA4* (Δ=0.111 mm^2^, p=0.011), *BEST1* (Δ=0.308 mm^2^, p=0.0012), and *PRPH2* (Δ=0.305 mm^2^, p=0.0051) all exhibiting more nasal EZ-loss, and *EFEMP1* (Δ=-0.311 mm^2^, p=0.018), *PRPF31* (Δ=0.442 mm^2^, p=0.0038), and *RP1* (Δ=0.375 mm^2^, p=0.0039) exhibiting greater temporal loss. *ABCA4* and *PRPH2* also exhibited significantly more nasal RPE-loss with Δ=0.191 mm^2^, p=1.3×10^-5^ and Δ=0.234 mm^2^, p=0.0084 respectively, and *EFEMP1* exhibited greater temporal loss, Δ=-0.296 mm^2^, p=0.0065.

### Genotype-phenotype correlations stratified by mode of inheritance

We analyze the phenotypes of genes associated with both recessive and dominant inheritance patterns in our data. In *BEST1* we see higher ICS and SRF in the recessive forms of the condition, which also results in higher retina thickness (**Figure 4a**). In *PROM1* we observe higher EZ- and RPE-loss overall in the recessive form whereas in the dominant form EZ- and RPE-loss are limited to the center (**Figure 4b**). In *RP1*, the recessive form has higher EZ- and RPE-loss and a decreased retina thickness (**Figure 4c**). In *GUCY2D,* the dominant form has increased central EZ- and RPE-loss, compared to the recessive form which is milder and has a thicker retina (**Figure 4d**). In *NR2E3*, the dominant form has widespread EZ-loss and increased inferior RPE-loss, whereas the recessive form is milder (**Figure 4e**).

**Figure 4:**
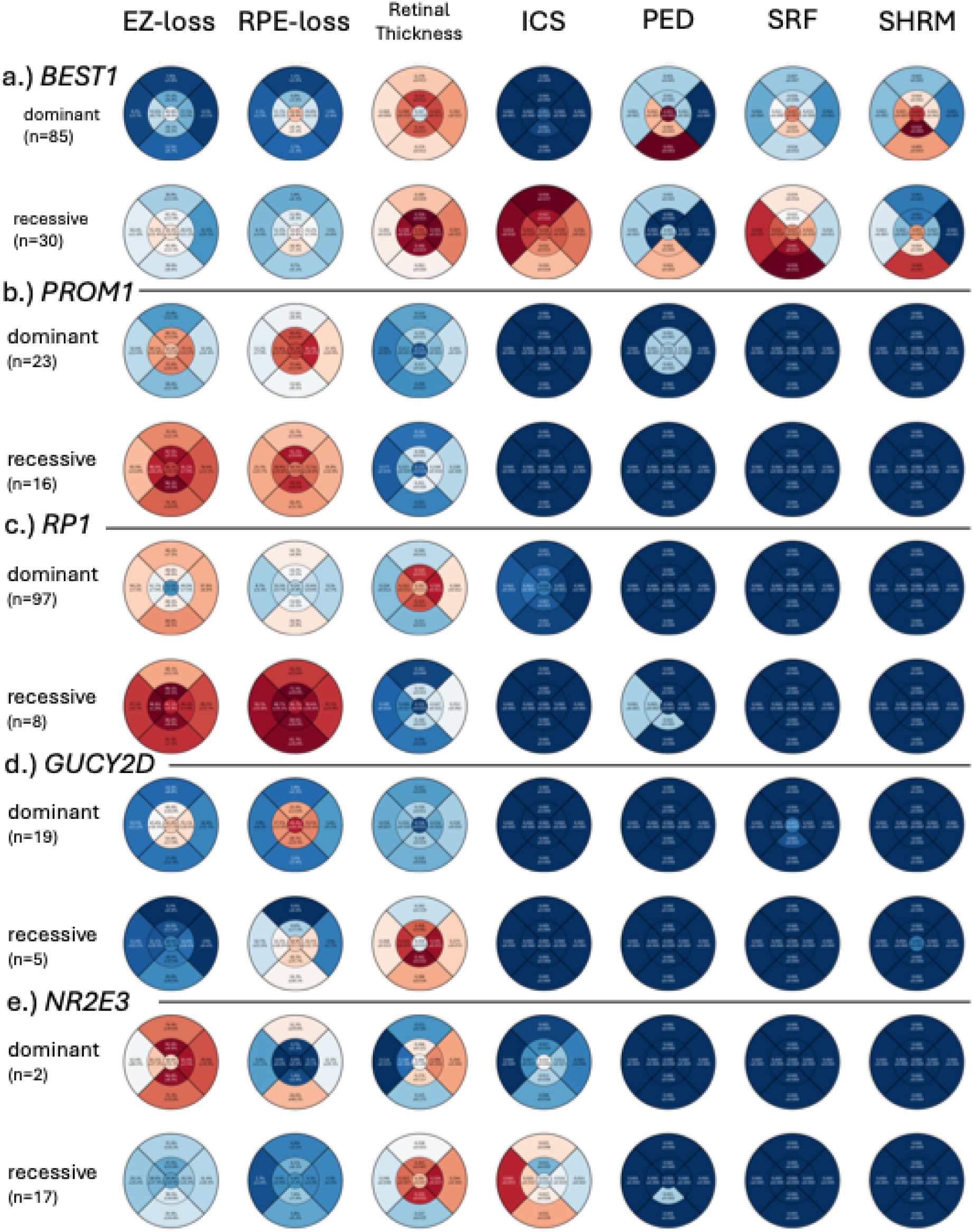
EDTRS region plots highlighting different phenotypes between dominant and recessive forms of disease for a.) *BEST1*, b.) *PROM1*, c.) *RP1*, d.) *GUCY2D* and e.) *NR2E3*. The recessive forms of disease are more severe for *BEST1*, *PROM1* and *RP1* and less severe for *GUCY2D* and *NR2E3*.

**Figure 5:**
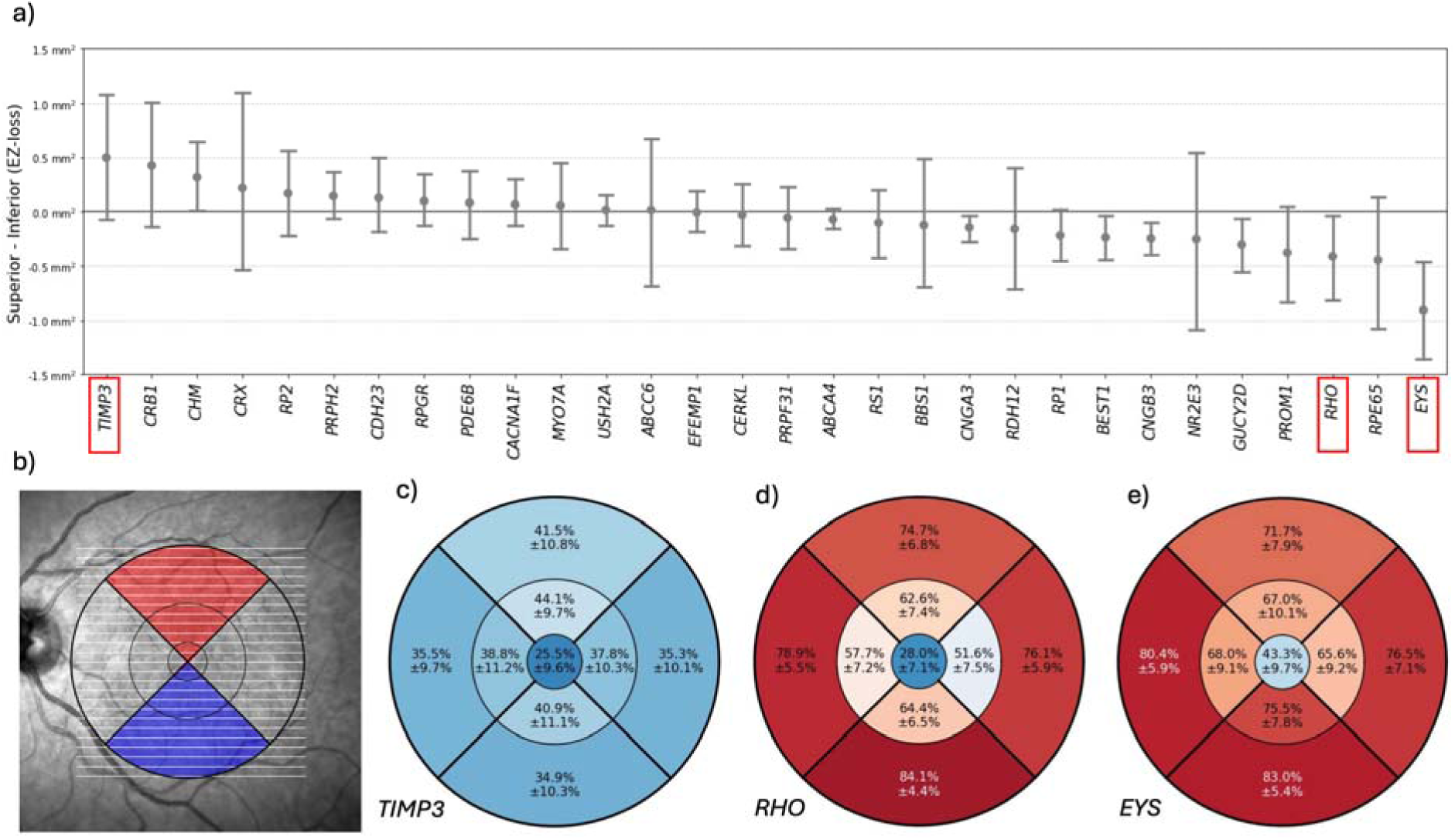
a) Per gene distribution of the difference between EZ-loss per SD-OCT volume in the superior and inferior quadrants **b)** Example projection of SD-OCT volume onto its corresponding IR enface with ETDRS sectors (1mm, 3mm, and 6mm diameter rings) and the projected areas of the full Superior (in red) and Inferior (in blue) quadrants. **c-e**) ETDRS segment plots with average occupancy and 95% confidence intervals for three genes **c)** *TIMP3* (Δ=0.496 mm^2^, p=0.11), **d)** *RHO* (Δ=-0.414 mm^2^, p=0.036) and **e)** *EYS* (Δ=-0.908 mm^2^, p=1.5×10^-4^).

### Genotype-phenotype sub-gene variant analysis

Comparing *USH2A* patients by number of truncating variants^29^, we find evidence for greater EZ-loss for patients with one or more truncating variants (**Figure 8b**). Patients with two truncating variants experiencing an average of 31.2±2.1 mm^2^ of EZ-loss, compared with 28.1±1.3 mm^2^ for patients with a single truncating variant, and 27.7±1.8 mm^2^ for patients with no truncating variants (p<9×10^-4^ one-way ANOVA). Though RPE-loss followed a similar trend for all patients with 5.9±1.8 mm^2^, 5.0±1.5 mm^2^, and 5.4±1.4 mm^2^ for patients with zero, one, and two truncating variants respectively, it was not significant (p<0.7 one-way ANOVA).

We also compare disease severity patients with variants in *RPGR* based on the region of the variation, specifically looking at the regions defined in Woof et al. ^20^(**Figure 8a)**. We find evidence for differences in EZ-loss by region with patients with variants with 27.0±1.8 mm^2^ for patients with a variants with alterations between the 600^th^ and 940^th^ amino acid, compared to 21.2±4.3 mm^2^ for patients with a variants with alterations at positions after the 940^th^ amino acid, and 29.1±1.5 mm^2^ for patients with positions before the 600^th^ amino acid (p<0.0012 one-way ANOVA). RPE-loss followed a similar pattern with 6.9±3.1 mm^2^, 8.2±1.9 mm^2^, and 5.0±2.7 mm^2^ for patients with variants >600, 600-940, and >940 respectively, but this was not significant (p=0.27).

In *PRPH2* we found that patients with the common missense mutations p.(Arg172Trp) and p.(Arg172Gln) were associated with significantly greater average EZ-loss (19.2±3.8 mm^2^ vs 9.4±2.1 mm^2^, p<2.3×10^-5^) and RPE-loss (11.2±3.5 mm^2^ vs 3.7±1.4 mm^2^, p<1.1×10^-5^) compared to patients with other variants (**Figure 8c**).

We applied AIRDetect-OCT to scans from patients affected by *ABCA4* associated retinopathy that were classified based on their genotype. Clinical severity of ABCA4 associated retinopathy has been associated/correlated with severity of genotype^30–34^. Patients were classified into four groups (A, B, C, and D) based on the severity of genetic variants as defined by Cornelis *et al.* ^35,36^. Patients in group A had two severe variants, while those in group C had a mild variant in trans with any other variant. Patients with variants of known severity whose combination did not fit the other two groups were placed into group B. We also included a fourth group (group D) of patients with the *ABCA4* allele p.(Gly1961Glu), which is known to be associated with a milder phenotype disease ^34,37–39^.

Patients in group D exhibited the greatest average retinal thickness, and the lowest area of EZ- and RPE-loss, in keeping with the milder phenotype (**Figure 6a**). Patients in group C showed the greatest volume of SHRM (**Figure 6b**). Overall, the area of EZ-loss and RPE-loss was related to the severity of genetic variants with group A showing the greatest area of loss (**Figure 6c**, **Figure 6d**).

**Figure 6:**
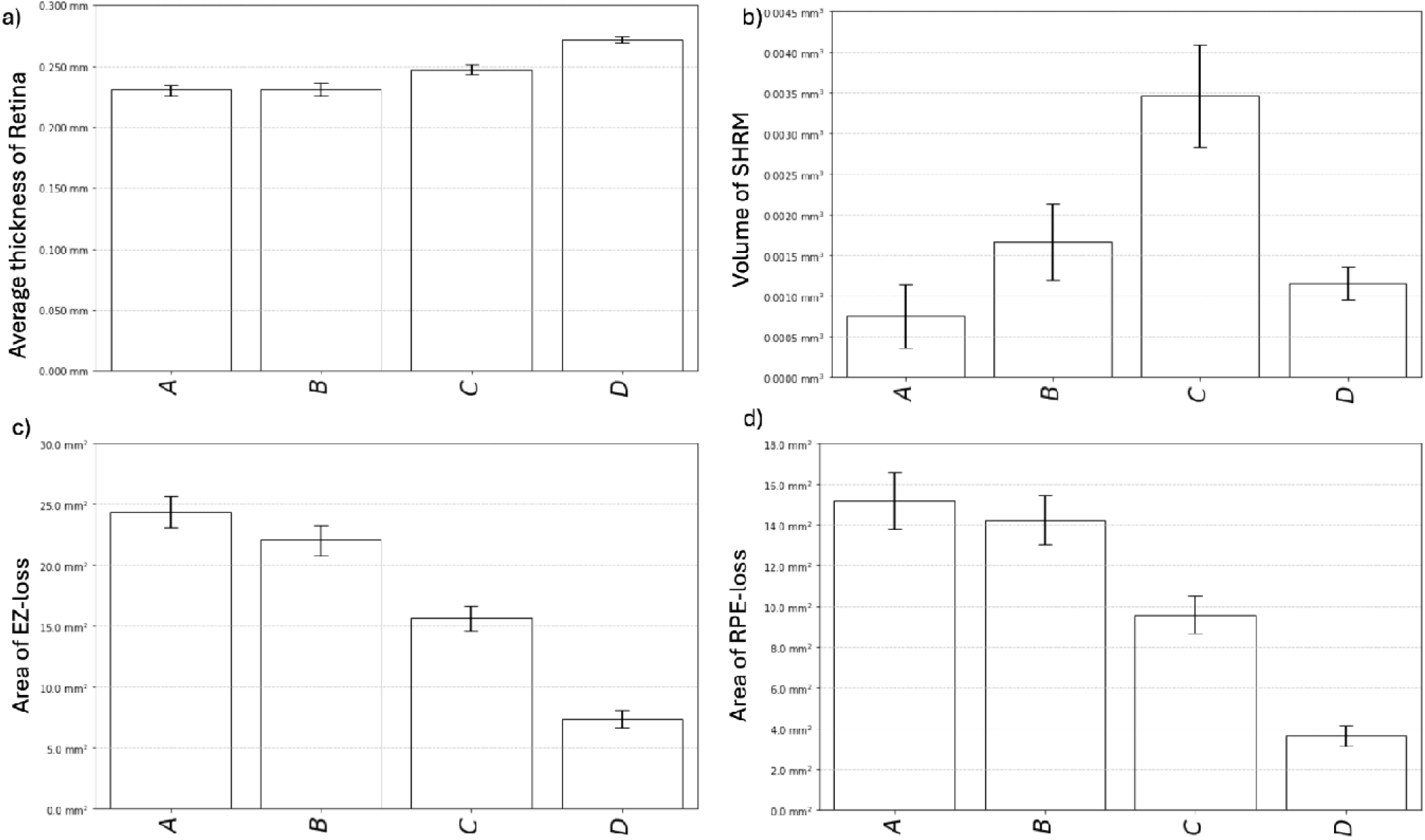
(**a**) Average retinal thickness, (**b**) volume of SHRM, (**c**) area of EZ-loss and (**d**) area of RPE-loss in the A, B, and C groups as defined by Cornelis et al.^35^ for *ABCA4* with the additional D group which consists of patients with p.(Gly1961Glu) variant.

### Longitudinal analysis in *ABCA4*

In addition to cross-sectional analysis by feature area we also looked at leveraging follow up appointments to calculate average rates of progression by patient (**Figure 7**). Analyzing the rate of disease progression by genotype group in *ABCA4* patients showed that severe variants were associated with significantly faster rates of progression (p<10^-4^). Patients in group A experienced the fastest rate of EZ-loss with an average rate of 2.80±0.62 mm^2^/yr, compared to 1.54 ±0.35 for group B, 1.42±0.33 for group C, and 0.56±0.18 for group D. Patients in group A also experienced a greater rate of RPE-loss at 1.67±0.80 mm^2^/yr, compared to 1.20±0.38, 1.13±0.31, and 0.73±0.29 for groups B, C, and D. It is clinically noteworthy that the rate of EZ loss was found to be greater than rate of RPE-loss, as has been shown previously^38^. There was also a significant difference (p=0.047) between the rate of RPE-loss between male and female patients in group A (**Table 6**). This was accompanied by substantial differences in patient numbers with female patients overrepresented compared to males in groups C and D, and underrepresented in group A (**Table S4**).

**Figure 7:**
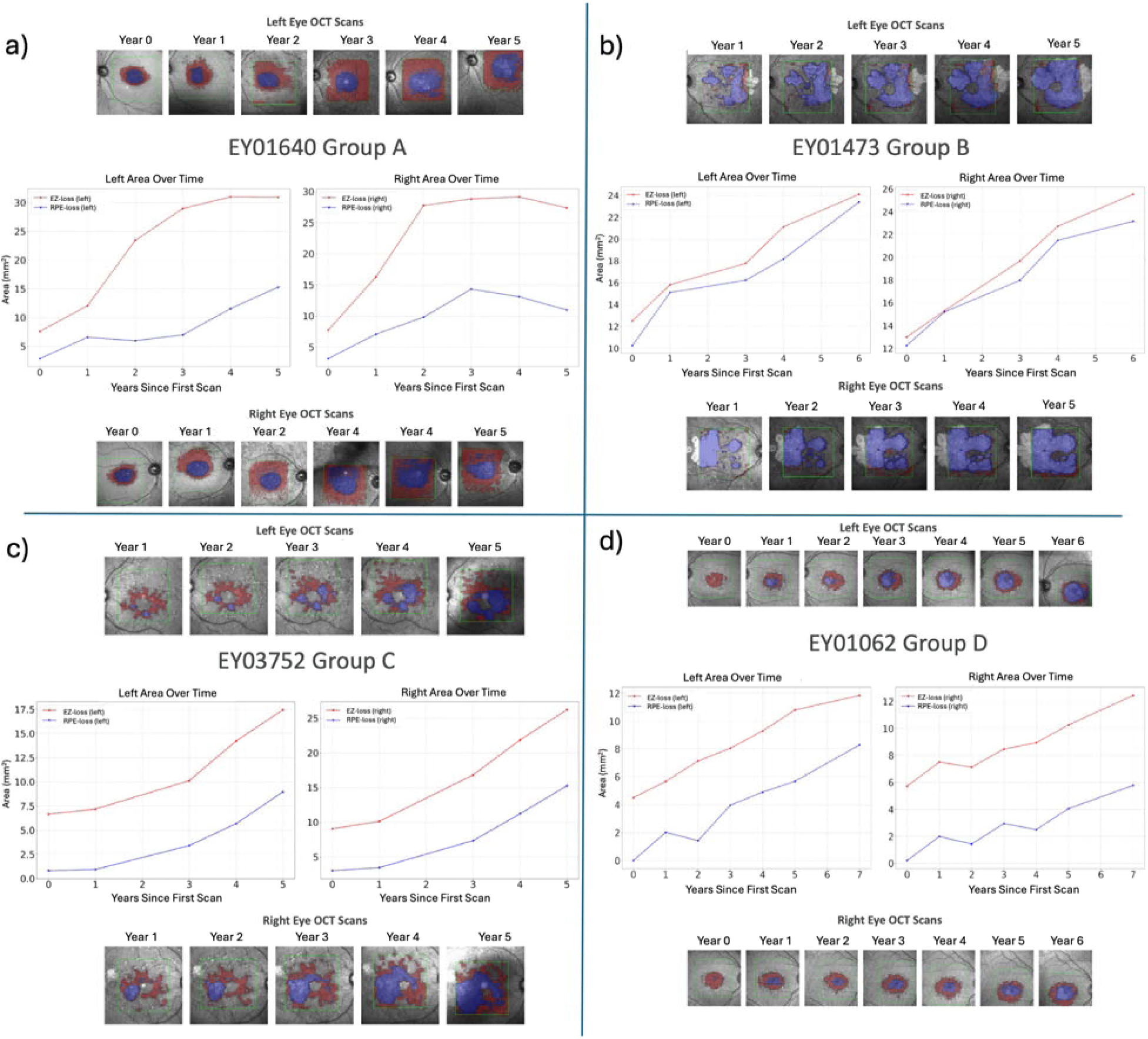
Progression plots of EZ-loss (red) and RPE-loss (blue) for the four genotypic groups as defined by Cornelis et al. with an additional group D consisting of patients with p.(Gly1961Glu) variant. Examples of ABCA4-retinopathy patients from (**a**) group A, (**b**) group B, (**c**) group C, (**d**) group D.

**Figure 8:**
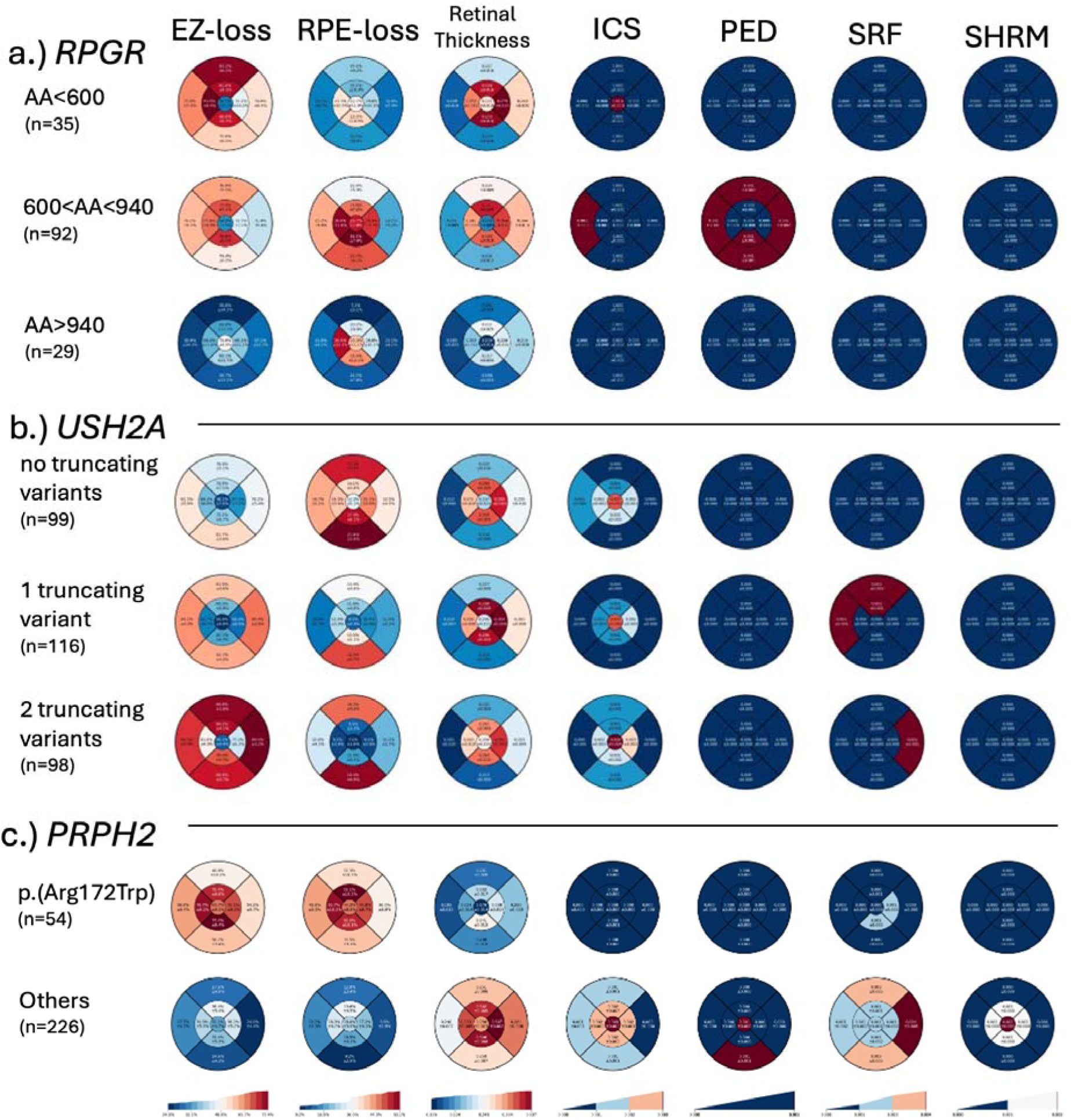
EDTRS region plots highlighting different phenotypes between different genotypes of a.) *RPGR*, comparing region of variation by amino acid sequence, b.) *USH2A*, comparing patients with zero, one, and two truncating variants, and c.) PRPH2, comparint patients with the common p.(Arg172Trp) variant to patients with other variants.

**Table 6:** Average per-patient rate of area of EZ- and RPE-loss for the four ABCA4 patient groups, disaggregated by patient sex (M=males, F=females). P-values are given for one-way ANOVA.

| Group | Age (mean) | EZ-loss (mm<sup>2</sup>/yr ±CI<sub>95</sub>) | | | RPE-loss (mm<sup>2</sup>/yr ±CI<sub>95</sub>) | | |
| --- | --- | --- | --- | --- | --- | --- | --- |
|  |  | All | M | F | All | M | F |
| A | 34.3 | 2.80 ±0.62 | 2.86 ±0.76 | 2.71 ±1.06 | 1.67 ±0.80 | 2.48 ±1.40 | 0.87 ±0.62 |
| B | 43.4 | 1.54 ±0.35 | 1.29 ±0.75 | 1.66 ±0.38 | 1.20 ±0.38 | 1.04 ±0.63 | 1.35 ±0.46 |
| C | 51.0 | 1.42 ±0.33 | 1.24 ±0.46 | 1.59 ±0.48 | 1.13 ±0.31 | 0.76 ±0.55 | 1.37 ±0.37 |
| D | 45.4 | 0.56 ±0.18 | 0.33 ±0.26 | 0.71 ±0.24 | 0.73 ±0.29 | 0.51 ±0.41 | 0.96 ±0.41 |
| p-value | 3.0×10<sup>-10</sup> | 1.2×10<sup>-13</sup> | 3.2×10<sup>-8</sup> | 6.4×10<sup>-6</sup> | 0.067 | 0.0060 | 0.31 |

## Discussion

This study presents the results of AIRDetect-OCT, the first AI algorithm to comprehensively segment eight clinically relevant features in SD-OCT images across all types of IRDs and to correlate those with demographics and genotypes. We have benchmarked AIRDetect-OCT against expert manual segmentation which showed high levels of model-grader agreement for the segmentation of features. We demonstrated the application of the algorithm to a produce a large real-world dataset of annotated scans, consisting of 7,405 OCT volumes from 3,534 patients with diseases associated with sequence variants in 176 different IRD genes. This identified consistent genotype-phenotype correlations and that the model can be used to automatically provide information on disease severity and progression using *ABCA4* associated retinopathy as an exemplar. AIRDetect-OCT provides a validated method for automatic feature segmentation that is important for quantifying disease severity, monitoring disease progression, providing information on prognosis to patients in the clinic, natural history studies, and clinical trial patient selection both in terms of most appropriate for intervention and accurately serially measuring the response to treatment.

A number of deep learning approaches for OCT segmentation in specific IRDs have been reported, however, the number of features segmented is limited and their generalisability to all types of IRDs has not been demonstrated. Loo *et al.* report a mean Dice of 79% for segmentation of the EZ on OCT using a deep learning model originally developed for macular telangiectasia Type 2 ^40^. A convolutional neural network (CNN) developed by Camino *et al.* for segmenting the preserved EZ on *en face* OCT in patients with choroideremia and RP achieved an accuracy of approximately 90% ^12^. Wang *et al.* applied a CNN to automatically identify the EZ on OCT in patients with RP and reported an accuracy of 91% ^13^. Wang et al. later developed a hybrid model composed of two CNNs for automatic segmentation of retinal layers on OCT in patients with RP^41^. They reported an overall improvement in accuracy with their hybrid model and EZ width highly correlated (r > 0.95) with manual segmentation^41^. The hybrid model underwent training with additional data by Wang *et al.* and this improved accuracy of the EZ segmentation with a reported Dice of 86.7%, and using deep learning with manual correction they were able to achieve a Dice of 85.24% ^14^. In our study, our reported accuracy and Dice for EZ-loss segmentation using AIRDetect-OCT is lower at 78.1% and 69% respectively. However in our study we use the same model for segmenting EZ loss in IRDs associated with 176 different genes and many different phenotypes which is a more challenging task.

Automatic segmentation of schitic cavities in patients with X-linked retinoschisis using deep learning was performed by Wei *et al.*^15^. Five deep learning approaches were benchmarked and their best results were a per-pixel accuracy of 99.3% and Dice of 85.7% with the U-Net++. The problem of automatically segmenting schitic cavities is the same as detecting ICS on OCT. In this study we achieved an accuracy of 93.6% and Dice of 89.2% in detecting ICS. Our model’s accuracy and model-grader agreement Dice were similar to that reported by Wei *et al* ^15^. Furthermore, our model was applied to a wide range of retinal phenotypes that were associated with 176 different genes.

Eckardt *et al.*^19^ also describe five different AI-based image segmentation models that were trained and tested in a small cohort of healthy controls and patients with IRDs (16 patients, 25 volume scans). They report achieving a Dice of 98.7% in their held-out IRD patient test set for retinal layer segmentation that can be used to provide retinal thickness. Kugelman *et al.* report the results of their fully semantic deep learning segmentation method and graph-search in patients with Stargardt disease. Their best performing semantic segmentation model achieved a mean absolute difference compared to the ground truth of 2.10 µm and 0.059mm^3^ for the retinal thickness and retinal volume, respectively^16^. Mishra et al. used a deep learning-shortest path framework to perform automated 11 retinal layer segmentation in patients with Stargardt disease and reported a mean absolute difference for ILM, outer RPE and Stargardt features of 0.77 ± 2.15, 1.55 ± 3.38 and 1.92 ± 3.71 pixels, respectively ^17^. In our study we report a model-grader agreement Dice of 95% for retinal segmentation, results that are comparable to those reported in the literature from previous studies with less heterogeneity within the IRD cohorts included.

The model-grader Dice for SRF and SHRM in our study was high at 92.3% and 85.7%, respectively, with a similarly high level of presence/absence accuracy of 95.8% and 92.2%, respectively. Though these were helped by relatively low rates of the respective features. The precision and recall for the detection of PED as a feature was 35.8% and 63.2%, respectively. This suggests that the model has a higher false positive rate, and tended to over-predict. The inter-grader Dice (84.7% and 77.4%), model-grader Dice (69.2% and 68.7%), and accuracy (78.1% and 76.4%) was lowest for EZ-loss and RPE-loss, respectively. The lower model-grader Dice was also reflected in the lower inter-grader Dice achieved from manual grading.

Performing genotype-phenotype correlations using AIRDetect-OCT showed that *RS1* and *NR2E3*, which are associated with macular schisis in X-linked retinoschisis and enhanced s-cone syndrome, respectively, were associated with higher volumes of ICS. *BEST1* was associated with the highest volume of SRF and SHRM as would be expected in patients with Best vitelliform macular dystrophy. *EFEMP1* was associated with the highest volume of PED/drusen and an increased volume of SHRM, a consistent feature in patients with autosomal dominant drusen. Retinal thickness was highest in *CRB1*, a characteristic feature of *CRB1*-associated retinopathy^42,43^. *NR2E3* and *RS1*, which are associated with ICS also showed higher average retinal thickness. Similarly, *BEST1* and *EFEMP1* associated retinal dystrophies, which are associated with higher volumes of SHRM and PED, respectively, were also associated with higher average retinal thickness, possibly due to vitelliform lesions above the Bruch’s membrane ^44,45^. Genotype-phenotype correlations in a subset of the 14 most common genes associated with RP in our cohort failed to demonstrate any clear trends in macular retinal thickness, EZ-loss or RPE-loss between genes with the same inheritance pattern at baseline. *RPGR* and *RP2*, which are associated with X-linked RP, were associated with lower volumes of ICS. X-linked RP is associated with a more severe phenotype compared to other forms of RP and usually presents in childhood ^46,47^. Reduced ICS would be expected to be associated with outer retinal degeneration affecting the macula, as would be seen in a more severe phenotype.

Using AIRDetect-OCT we were able to show that in patients with *ABCA4* associated retinopathy more severe genotypes were associated with higher areas of EZ-loss and RPE-loss at baseline. In doing so we observed substantial differences in severity between male and female patients, particularly for the more severe groups A and B, where male patients in these groups exhibited greater EZ- and RPE-loss. Sex disparities were also present in the number of patients in each category, which could indicate differences in disease penetrance. Analyzing rate of disease progression based on EZ-loss and RPE-loss we similarly demonstrated a trend with more severe genotypes associated with faster rates of disease progression. However, our results did not identify a statistically significant difference when comparing average retinal thickness, area of EZ-loss, and area of RPE-loss between groups A and B. Missense variants in *ABCA4* have been seen more frequently than expected in patients with severe phenotypes^37^ and *in vitro* functional analysis of missense variants has classified some of these as severe. The classification of variants based on severity may have under classified patients with severe missense variants into group B thereby masking any true difference in average retinal thickness, area of EZ-loss, and area of RPE-loss between these two groups with the most severe variants.

Limitations of our study include phenotypic variability. For example, *NR2E3* being predominantly associated with enhanced S cone syndrome in autosomal recessive cases and autosomal dominant RP associated with the p.(Gly56Arg). Patient age is also a big confounder, hence the reported averages over a patient cohort are heavily dependent on age mix. This study was carried out using real-world data which means that there was considerable variability in the quality and parameters of the images, especially in cases with more advanced disease where it can be more difficult to acquire images. Advanced disease where the RPE is lost complicates the identification of features such as distinguishing SHRM, which lies above the RPE, from PED, which lies below the RPE. In terms of algorithm performance, we found that recall was higher than precision for all features, suggesting that the model tended to overpredict rather than underpredict (**Table 2**). We also found that PED and SHRM were challenging for the algorithm to distinguish, likely due to the RPE frequently being disrupted in these conditions, complicating the localisation of these features relative to the RPE, hence they were often confused (**Table S2**). This explains the lower precision for PED of 36% in **Table 2**. Combining PED and SHRM into one class improved the performance of PED to 94.6% accuracy, 66.0% precision and 78.0% recall. Additionally, AIRDetect was trained to detect the absence of EZ and the RPE, which precludes the measurement of EZ-position, and thickness/attenuation which can be important biomarkers in certain conditions^18^. Predicting only presence/absence of the EZ was felt to be advantageous to segmenting the EZ layer directly, due to the inherent difficulties in segmenting the EZ. However future work may involve introducing a direct EZ segmentation model to enable measurement of the thickness of the EZ. Our study was also limited to macular SD-OCT scans covering the central 20-degree portion of the retina. In future, we hope to expand the capabilities of AIRDetect to work with wide-angle SD-OCT to enhance applicability to early-stage RP cases.

Our model’s performance highlights that automatic segmentation can identify specific biomarkers that are associated with IRDs. Correlation between the structure and function based on the features identified on OCT images will enable a deeper understanding of the natural history of different IRD conditions. Automatic segmentation will likely become increasingly important in optimizing clinical trial design and clinical management.

Future work will involve image registration using vasculature key points across timepoints to enhance accuracy of longitudinal analysis and co-registration of structure with functional assessments.

## Data Availability

The data that support the findings of this study are divided into two groups, published data and restricted data. Published data are available from the Github repository. Restricted data are curated for AIRDetect-OCT users under a license and cannot be published, to protect patient privacy and intellectual property. Synthetic data derived from the test data has been made available at https://github.com/Eye2Gene/.

https://www.eye2gene.com

## Ethics

This research was approved by the IRB and the UK Health Research Authority Research 403 Ethics Committee (REC) reference (22/WA/0049) “Eye2Gene: accelerating the diagnosis of 404 inherited retinal diseases” Integrated Research Application System (IRAS) (project ID: 405 242050). All research adhered to the tenets of the Declaration of Helsinki.

## Code availability

The source code for the AIRDetect-OCT model architecture training and inference is available from https://github.com/Eye2Gene/. The model weights of AIRDetect-OCT are intellectual proprietary of UCLB so cannot be shared publicly. However, they may be shared via a licensing agreement with UCLB. A running online version of the AIRDetect-OCT app is accessible via the Eye2Gene website (www.eye2gene.com) on invitation.

## Author contributions

WAW analyzed the data and wrote the manuscript. NP obtained the funding, designed the experiments, analyzed data and wrote the manuscript. MiM, KB, WAW, TACG, SAK, MDV, BM designed the experiments, analyzed data and wrote the manuscript. SS analyzed the data. MS wrote the manuscript. PBa, PBu, DGP, YL analyzed the data. All authors, WAW, TACG, SAK, MDV, SS, PBa, BSM, MS, PBu, DGP, SL, GN, ASS, BG, BL, DJF, MG, YL, QN, YFK, DS, JF, PJP, IM, MaM, JS, SRDS, BL, FGH, KF, ARW, OAM, SMD, SM, KB, MiM and NP, have contributed to the writing and critically reviewed the manuscript.

## Acknowledgement

This work is primarily funded by a NIHR AI Award (AI_AWARD02488) which supported NP, WAW, MM, KB, SD and SM. The research was also supported by a grant from the National Institute for Health Research (NIHR) Biomedical Research Centre (BRC) at Moorfields Eye Hospital NHS Foundation Trust and UCL Institute of Ophthalmology. NP was also previously funded by Moorfields Eye Charity Career Development Award (R190031A). BJ was partially funded by IIR-DE-002818 from Shire/Takeda and by the European Reference Network for Rare Malformation Syndromes, Intellectual and Other Neurodevelopmental Disorders (ERN-ITHACA). OAM is supported by the Wellcome Trust (206619/Z/17/Z). AYL is supported by an unrestricted and career development award from RPB, Latham Vision Science Awards, NIH OT2OD032644, NEI/NIH K23EY029246, and NIA/NIH U19AG066567. SA is supported by a scholarship from Qatar National Research Fund (GSRA6-1-0329-19010). SL is supported by a MRC Clinician Scientist Fellowship. This project was also supported by a generous donation by Stephen and Elizabeth Archer in memory of Marion Woods. The hardware used for analysis was supported by the BRC Challenge Fund (BRC3_027). We also gratefully acknowledge the support of NVIDIA Corporation with the donation of the Titan Xp GPU used for this research. The views expressed are those of the authors and not the funding organizations.

## Supplementary

**Figure S1:**
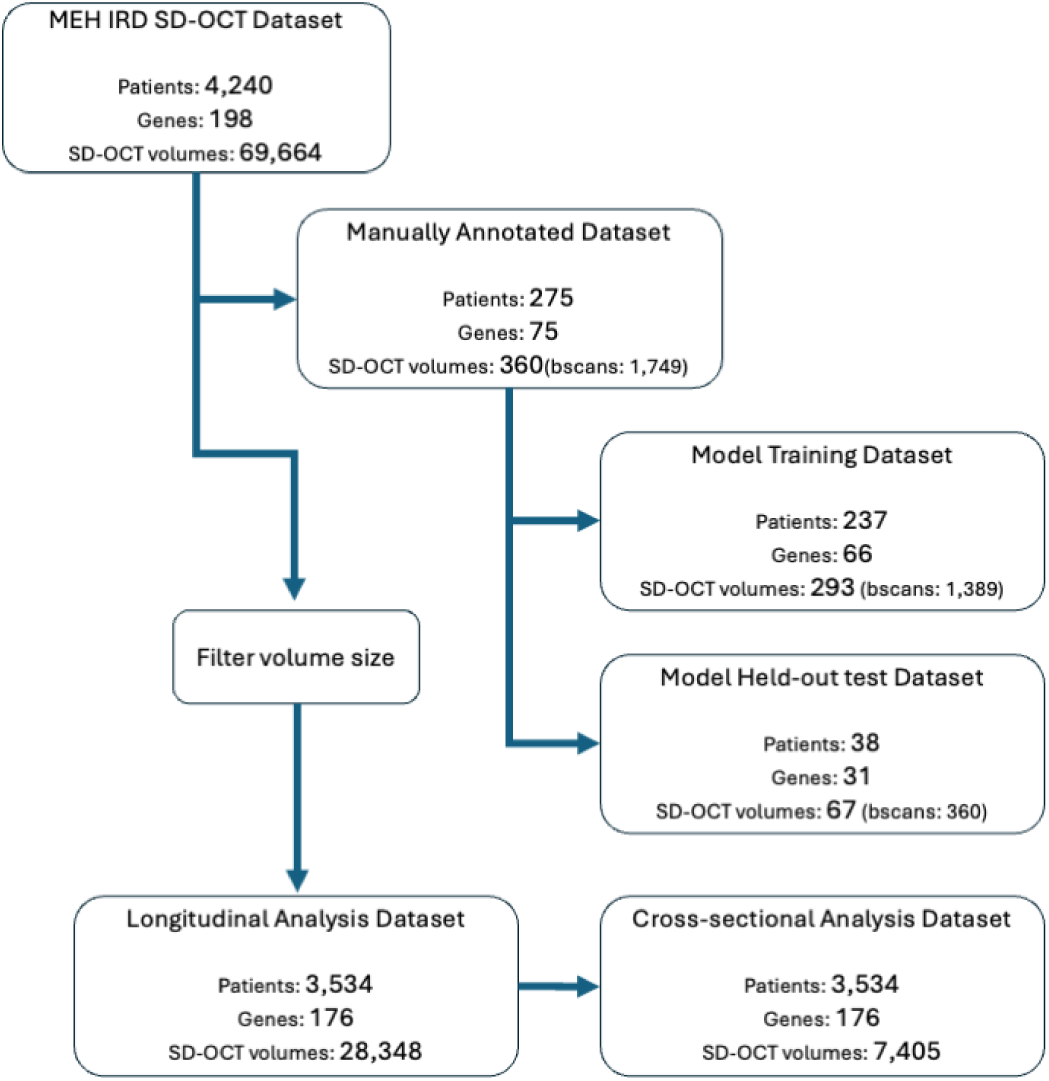
Data flow indicating how many patients and corresponding SD-OCT volumes were used to train and test AIRDetect-OCT. The cross-sectional analysis data was produced by filtering by enface dimensions and number of b-scans, only the volume at baseline was kept. For the longitudinal analysis dataset all follow up volumes were included.

**Table S1:** Mean value and 95% confidence interval (defined as standard error * 1.96) of annotations per gene for the 30 most common genes. Annotations include: Retina, Intraretinal Cystic Spaces (ICS), Subretinal Fluid (SRF), Subretinal Hyper-reflective Material (SHRM), Pigment Epithelium Detachment (PED), Ellipsoid Zone Loss (EZ-loss) and Retinal Pigment Epithelium Loss (RPE-loss).

|  |  |  | Retina Thickness (mm) |  | ICS Volume (mm<sup>3</sup>) |  | SRF Volume (mm<sup>3</sup>) |  | SHRM Volume (mm<sup>3</sup>) |  | PED Volume (mm<sup>3</sup>) |  | EZ-loss Area (mm<sup>2</sup>) |  | RPE-loss Area (mm<sup>2</sup>) |  |
| --- | --- | --- | --- | --- | --- | --- | --- | --- | --- | --- | --- | --- | --- | --- | --- | --- |
| gene | patients | age | Mean | CI 95% | Mean | CI 95% | Mean | CI 95% | Mean | CI 95% | Mean | CI 95% | Mean | CI 95% | Mean | CI 95% |
| ABCA4 | 824 | 46.6 | 0.248 | 0.003 | 0.0018 | 0.0011 | 0.0015 | 0.0008 | 0.0023 | 0.0005 | 0.0056 | 0.0014 | 16.7 | 0.9 | 10 | 0.7 |
| ABCC6 | 18 | 61.9 | 0.261 | 0.021 | 0.0049 | 0.0047 | 0.016 | 0.0107 | 0.0061 | 0.0033 | 0.0458 | 0.0466 | 15 | 5 | 8.9 | 4 |
| BBS1 | 28 | 44.2 | 0.245 | 0.012 | 0.0002 | 0.0001 | 0.0055 | 0.0105 | 0 | 0 | 0.0008 | 0.0003 | 26.5 | 4.9 | 10.9 | 4.4 |
| BEST1 | 132 | 45.2 | 0.321 | 0.006 | 0.0914 | 0.0535 | 0.2071 | 0.0555 | 0.0448 | 0.0108 | 0.0164 | 0.0056 | 6.7 | 1.3 | 1.6 | 0.5 |
| CACNA1F | 27 | 26.2 | 0.275 | 0.009 | 0.0007 | 0.0013 | 0 | 0 | 0 | 0 | 0 | 0 | 2.4 | 1.8 | 0.4 | 0.4 |
| CDH23 | 21 | 38.2 | 0.283 | 0.017 | 0.0082 | 0.0083 | 0.0028 | 0.0054 | 0.0001 | 0.0001 | 0.0006 | 0.0007 | 21.1 | 6.4 | 5.9 | 3.9 |
| CERKL | 20 | 38.3 | 0.244 | 0.017 | 0.0003 | 0.0002 | 0 | 0 | 0 | 0 | 0.0004 | 0.0002 | 33.2 | 1.9 | 7.1 | 3.3 |
| CHM | 108 | 44.3 | 0.265 | 0.01 | 0.0061 | 0.0036 | 0.0058 | 0.0077 | 0.0004 | 0.0005 | 0.0026 | 0.0019 | 20.9 | 2.1 | 15.2 | 1.9 |
| CNGA3 | 47 | 31.7 | 0.274 | 0.009 | 0.0002 | 0.0003 | 0.0008 | 0.0006 | 0.0018 | 0.0035 | 0.0149 | 0.0289 | 2.7 | 1.7 | 0.7 | 0.5 |
| CNGB3 | 64 | 36.0 | 0.274 | 0.005 | 0.0002 | 0.0001 | 0.0008 | 0.0008 | 0.0001 | 0.0001 | 0.0001 | 0.0001 | 3.1 | 1.2 | 0.5 | 0.3 |
| CRB1 | 44 | 35.4 | 0.334 | 0.014 | 0.0686 | 0.0563 | 0.0051 | 0.0098 | 0 | 0 | 0.0017 | 0.001 | 25.6 | 3.3 | 13.1 | 3.2 |
| CRX | 25 | 61.3 | 0.252 | 0.012 | 0.0024 | 0.0042 | 0 | 0 | 0 | 0 | 0.0004 | 0.0002 | 15.2 | 4.6 | 4.4 | 2.5 |
| EFEMP1 | 30 | 56.8 | 0.303 | 0.011 | 0.0154 | 0.018 | 0.0066 | 0.0062 | 0.0393 | 0.0241 | 0.3431 | 0.1221 | 4.1 | 1.7 | 2.3 | 1.2 |
| EYS | 69 | 55.1 | 0.256 | 0.009 | 0.0165 | 0.0213 | 0 | 0 | 0.0001 | 0.0001 | 0.0024 | 0.0025 | 27.2 | 1.9 | 6.7 | 2.1 |
| GUCY2D | 26 | 44.9 | 0.262 | 0.014 | 0.0001 | 0.0001 | 0.0046 | 0.0067 | 0.0003 | 0.0004 | 0.0005 | 0.0004 | 9.7 | 4.3 | 3.6 | 2 |
| MYO7A | 52 | 44.5 | 0.287 | 0.012 | 0.0184 | 0.0162 | 0.0001 | 0.0001 | 0 | 0 | 0.0016 | 0.0016 | 22.6 | 3.7 | 9.9 | 2.9 |
| NR2E3 | 26 | 44.4 | 0.324 | 0.023 | 0.2534 | 0.2598 | 0.0022 | 0.004 | 0.0016 | 0.0028 | 0.0052 | 0.0081 | 17.3 | 4.3 | 4.1 | 3.1 |
| PDE6B | 30 | 44.1 | 0.258 | 0.011 | 0.0392 | 0.0255 | 0 | 0.0001 | 0 | 0 | 0.0013 | 0.0022 | 30.8 | 1.3 | 8.8 | 3.7 |
| PROM1 | 46 | 44.9 | 0.252 | 0.01 | 0.0001 | 0.0001 | 0 | 0.0001 | 0 | 0 | 0.0026 | 0.0021 | 21.3 | 3.6 | 8.1 | 2.9 |
| PRPF31 | 65 | 46.4 | 0.277 | 0.007 | 0.0324 | 0.0166 | 0.0004 | 0.0006 | 0 | 0 | 0.0004 | 0.0002 | 26 | 2.2 | 4.7 | 2 |
| PRPH2 | 146 | 58.7 | 0.278 | 0.007 | 0.0091 | 0.0091 | 0.0152 | 0.0266 | 0.0051 | 0.0019 | 0.0035 | 0.0013 | 11.7 | 2 | 5.5 | 1.5 |
| RDH12 | 25 | 35.6 | 0.276 | 0.016 | 0.0116 | 0.0144 | 0.0038 | 0.0053 | 0 | 0 | 0.0085 | 0.0066 | 27.5 | 2.3 | 15.5 | 3.7 |
| RHO | 104 | 48.1 | 0.265 | 0.006 | 0.0346 | 0.0166 | 0.0015 | 0.0028 | 0.0001 | 0.0001 | 0.0022 | 0.0035 | 27.1 | 1.6 | 5.6 | 1.7 |
| RP1 | 109 | 59.6 | 0.274 | 0.007 | 0.0119 | 0.008 | 0.0003 | 0.0005 | 0 | 0 | 0.0009 | 0.0007 | 23.6 | 2 | 5.8 | 1.7 |
| RP2 | 26 | 32.7 | 0.239 | 0.016 | 0 | 0 | 0 | 0 | 0 | 0 | 0.0004 | 0.0002 | 31.8 | 2.8 | 7.2 | 3.8 |
| RPE65 | 29 | 35.0 | 0.255 | 0.012 | 0.0001 | 0.0001 | 0 | 0 | 0.0027 | 0.0052 | 0.0011 | 0.0009 | 23.4 | 4.4 | 9.4 | 4.1 |
| RPGR | 173 | 41.7 | 0.25 | 0.005 | 0.002 | 0.0015 | 0.0001 | 0.0002 | 0.0001 | 0.0002 | 0.0024 | 0.0031 | 26.1 | 1.4 | 7.1 | 1.4 |
| RS1 | 105 | 33.8 | 0.31 | 0.012 | 0.4722 | 0.1378 | 0.0002 | 0.0002 | 0 | 0.0001 | 0.0018 | 0.0016 | 7.4 | 1.9 | 2.4 | 0.9 |
| *TIMP3* | 34 | 56.3 | 0.284 | 0.011 | 0.0048 | 0.0062 | 0.0071 | 0.0084 | 0.01 | 0.007 | 0.0446 | 0.0278 | 11.8 | 3.4 | 5.7 | 2.4 |
| *USH2A* | 319 | 50.6 | 0.26 | 0.003 | 0.0168 | 0.0061 | 0.0003 | 0.0005 | 0 | 0 | 0.0004 | 0.0001 | 29 | 0.8 | 5.4 | 0.9 |
|  | Mean | 43.7 | 0.262 | 0.001 | 0.0272 | 0.002 | 0.0104 | 0.0011 | 0.0027 | 0.0002 | 0.0064 | 0.0005 | 19.6 | 0.2 | 7.2 | 0.1 |

**Table S2:** Pixel-level confusion matrix of intraretinal Cystic Spaces (ICS), Subretinal Fluid (SRF), Subretinal Hyper-reflective Material (SHRM), Pigment Epithelium Detachment (PED) between manual (grader) and automatic (aIRDetect) annotations. Rows represent ground truth (manual annotation) and columns represent predictions (automatic annotations). Values are given as a proportion of ground truth label captured by that row (e.g. 4.12% of ICS area was incorrectly identified as SRF). PED and SHRM are more frequently confused for each other than the other features. This is to be expected given they are of similar pixel intensity and distinguishable by their position relative to the RPE, which is frequently absent in IRDs, leading to ambiguity.

|  | ICS | SRF | SHRM | PED |
| --- | --- | --- | --- | --- |
| ICS | 95.88% | 4.12% | 0.00% | 0.00% |
| SRF | 1.07% | 98.44% | 0.20% | 0.29% |
| SHRM | 0.02% | 4.06% | 77.37% | 18.56% |
| PED | 0.00% | 0.00% | 14.44% | 85.56% |

**Table S3:** Number of b-scans per volume after filtering by enface dimensions.

| Number of b-scans in volume | Count | % |
| --- | --- | --- |
| 25 | 4009 | 54.1% |
| 49 | 1777 | 24.0% |
| 19 | 1063 | 14.4% |
| 193 | 253 | 3.4% |
| 97 | 93 | 1.3% |
| 37 | 45 | 0.6% |
| 6 | 34 | 0.5% |
| All others | 131 | 1.8% |

**Figure S2:**
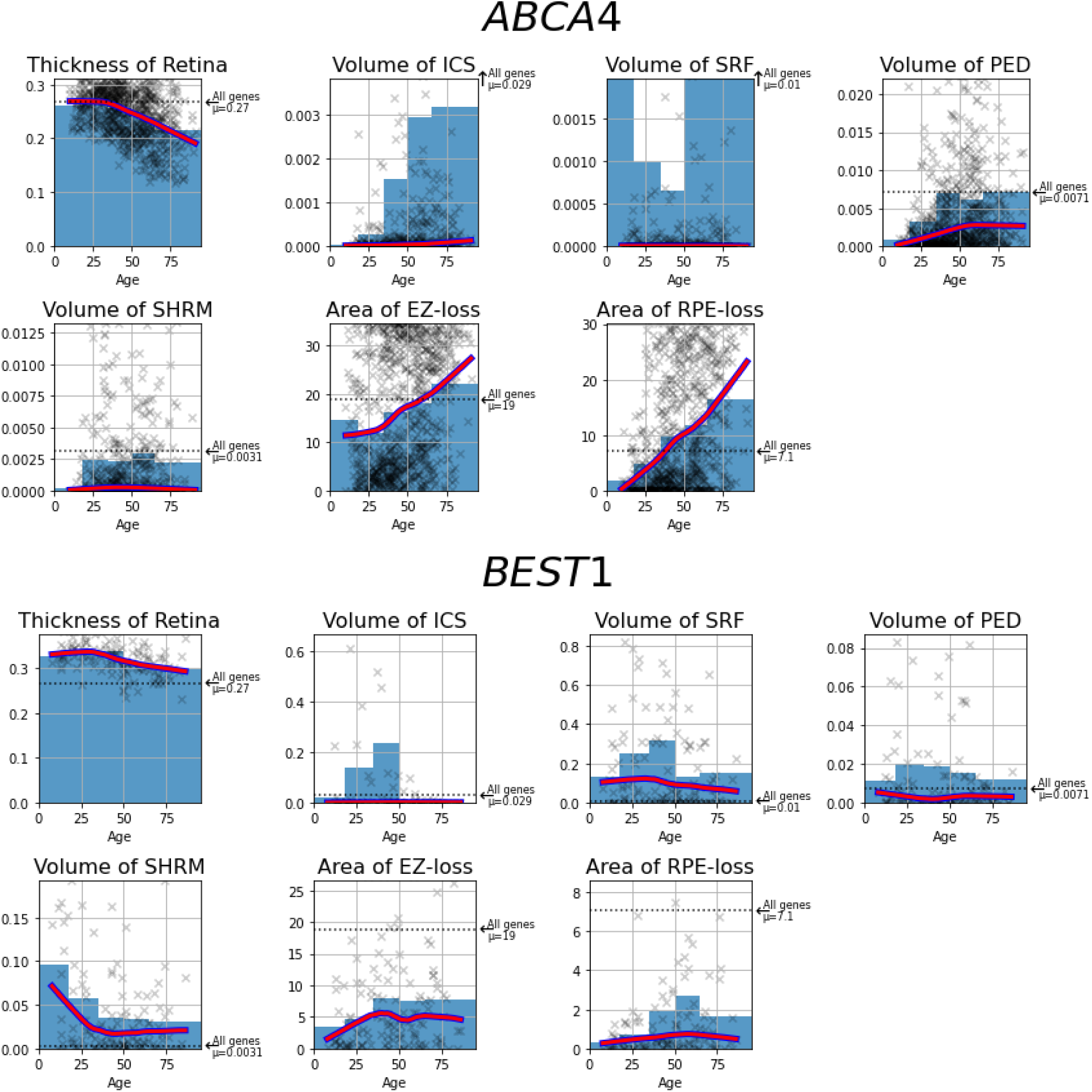
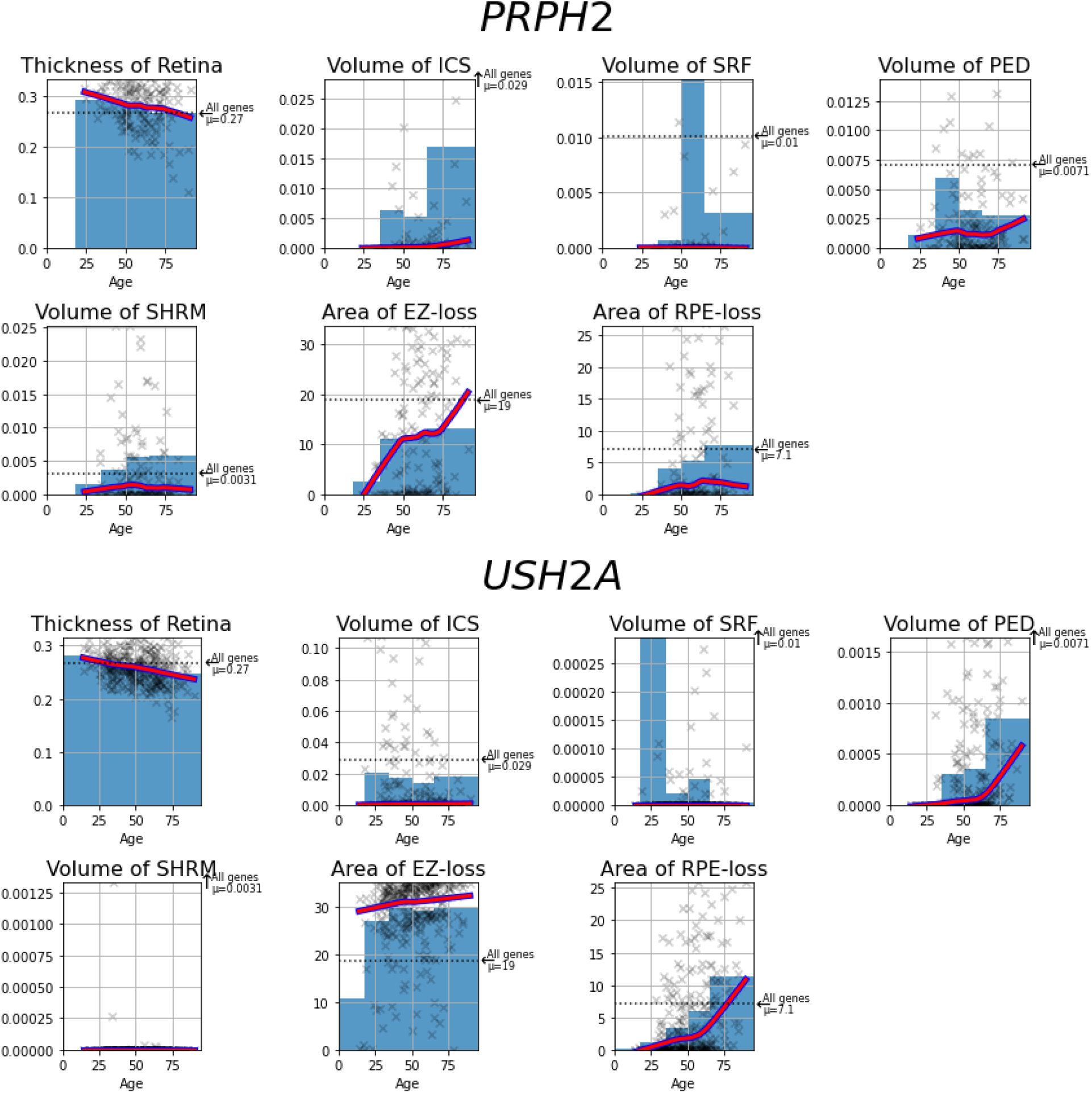
Effect of patient age on AIRDetect extracted retinal features for 5 select genes. Each point represents a single patient at their initial presentation, the red line is a LOWESS curve demonstrating the overall trend, the shaded region denotes the mean value for each age cohort (0-18, 19-35, 36-50, 51-65, and 66+), and the dotted line represents the mean value for the feature across all genes for comparison.

**Table S4:** Number of patients (n), median age, average per-patient area of EZ-loss, and RPE-loss for the four ABCA4 patient groups, split by patient sex.

| Group | n |  | Median Age |  | EZ-loss (mm<sup>2</sup> ±CI<sub>95</sub>) |  | RPE-Loss (mm<sup>2</sup> ±CI<sub>95</sub>) |  |
| --- | --- | --- | --- | --- | --- | --- | --- | --- |
|  | M | F | M | F | M | F | M | F |
| A | 51 | 25 | 30 | 28 | 25.8 ±2.9 | 21.5 ±4.2 | 17.3 ±3.4 | 11.0 ±3.7 |
| B | 50 | 45 | 41 | 41 | 24.2 ±3.4 | **19.7 ±3.4** | 16.1 ±3.2 | 12.2 ±3.3 |
| C | 57 | 73 | 53 | 51 | 16.8 ±3.1 | **14.7 ±2.7** | 11.1 ±3.0 | 8.4 ±2.2 |
| D | 56 | 73 | 41 | 42 | 7.7 ±2.3 | 7.2 ±1.7 | 4.1 ±1.7 | 3.3 ±1.2 |

**Table S6:**
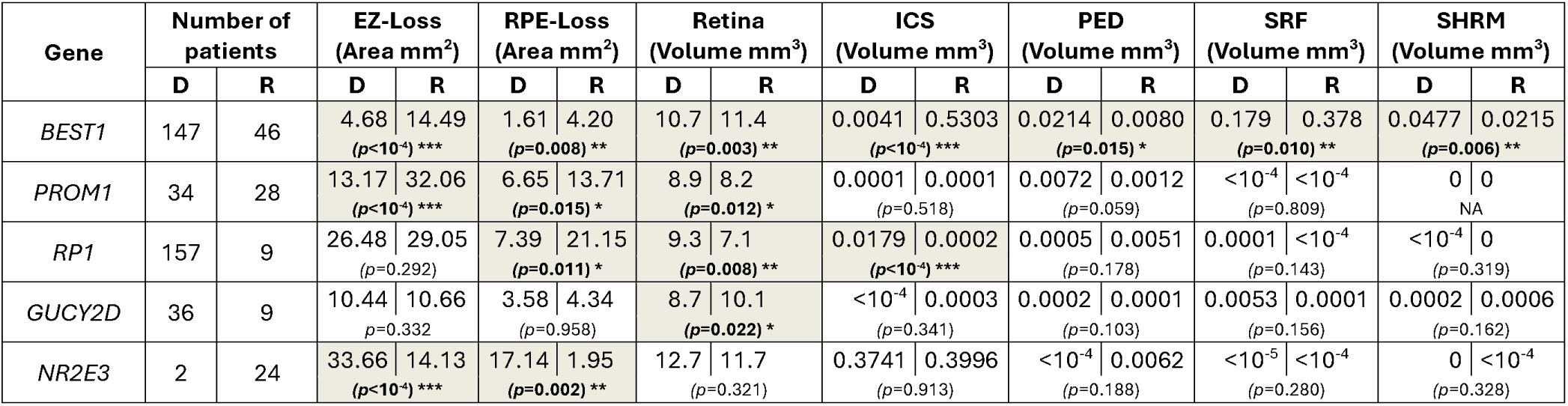
Mean values of ellipsoid zone loss (EZ-loss), retinal pigment epithelium loss (RPE-loss), retinal thickness (Retina), intraretinal cystic spaces (ICS), pigment epithelium detachment (PED), subretinal fluid (SRF) and subretinal hyper-reflective material (SHRM) for patients with dominant and recessive *BEST1*, *PROM1*, *RP1*, *GUCY2D* and *NR2E3*. Area and Volume are defined as averaged en-face area and volume respectively, within the 6mm diameter ring centred on the fovea. For each gene, the upper row presents the mean values for each feature, while the lower row displays the corresponding *p*-values using t-test (dominant vs recessive). D = dominant, R = recessive.

## References

1. Liew G, Michaelides M, Bunce C. A comparison of the causes of blindness certifications in England and Wales in working age adults (16–64 years), 1999–2000 with 2009–2010. BMJ Open 2014;4:e004015. Available at: [Accessed December 4, 2017].

2. Heath Jeffery RC, Mukhtar SA, McAllister IL, et al. Inherited retinal diseases are the most common cause of blindness in the working-age population in Australia. Ophthalmic Genet 2021;42:431–439.

3. Pontikos N, Wagner SK, Balaskas K, et al. Comment on: Trends in retina specialist imaging utilization from 2012 to 2016 in the United States medicare fee-for-service population. Am J Ophthalmol 2020;211:229.

4. Balaratnasingam C, Messinger JD, Sloan KR, et al. Histologic and optical coherence tomographic correlates in drusenoid pigment epithelium detachment in age-related macular degeneration. Ophthalmology 2017;124:644–656.

5. Al-Khuzaei S, Broadgate S, Foster CR, et al. An overview of the genetics of ABCA4 retinopathies, an evolving story. Genes (Basel) 2021;12:1241.

6. Al-Khuzaei S, Broadgate S, Halford S, et al. Novel pathogenic sequence variants in NR2E3 and clinical findings in three patients. Genes (Basel) 2020;11:1288.

7. Daich Varela M, Duignan ES, De Silva SR, et al. CERKL-associated retinal dystrophy: Genetics, phenotype, and natural history. Ophthalmol Retina 2023;7:918–931.

8. Hahn LC, Georgiou M, Almushattat H, et al. The Natural History of Leber Congenital Amaurosis and Cone-Rod Dystrophy Associated with Variants in the GUCY2D Gene. Ophthalmol Retina 2022;6:711–722.

9. Kiraly P, Cottriall CL, Taylor LJ, et al. Outcomes and Adverse Effects of Voretigene Neparvovec Treatment for Biallelic RPE65-Mediated Inherited Retinal Dystrophies in a Cohort of Patients from a Single Center. Biomolecules 2023;13. Available at: 10.3390/biom13101484.

10. Fahim AT, Bouzia Z, Branham KH, et al. Detailed clinical characterisation, unique features and natural history of autosomal recessive RDH12-associated retinal degeneration. Br J Ophthalmol 2019;103:1789–1796.

11. Bouzia Z, Georgiou M, Hull S, et al. GUCY2D-Associated Leber Congenital Amaurosis: A Retrospective Natural History Study in Preparation for Trials of Novel Therapies. Am J Ophthalmol 2019. Available at: http://www.sciencedirect.com/science/article/pii/S000293941930515X.

12. Camino A, Wang Z, Wang J, et al. Deep learning for the segmentation of preserved photoreceptors on en face optical coherence tomography in two inherited retinal diseases. Biomed Opt Express 2018;9:3092–3105.

13. Wang Y-Z, Galles D, Klein M, et al. Application of a deep machine learning model for automatic measurement of EZ width in SD-OCT images of RP. Transl Vis Sci Technol 2020;9:15.

14. Wang Y-Z, Juroch K, Birch DG. Deep learning-assisted measurements of photoreceptor ellipsoid zone area and outer segment volume as biomarkers for retinitis pigmentosa. Bioengineering (Basel) 2023;10. Available at: 10.3390/bioengineering10121394.

15. Wei X, Li H, Zhu T, et al. Deep learning with automatic data augmentation for segmenting schisis cavities in the optical coherence tomography images of X-linked juvenile retinoschisis patients. Diagnostics (Basel) 2023;13. Available at: 10.3390/diagnostics13193035.

16. Kugelman J, Alonso-Caneiro D, Chen Y, et al. Retinal boundary segmentation in Stargardt disease optical coherence tomography images using automated deep learning. Transl Vis Sci Technol 2020;9:12.

17. Mishra Z, Wang Z, Sadda SR, Hu Z. Automatic Segmentation in Multiple OCT Layers For Stargardt Disease Characterization Via Deep Learning. Transl Vis Sci Technol 2021;10:24.

18. Pinedo-Diaz G, Lorenz B, Künzel SH, et al. Deep learning-based SD-OCT layer segmentation quantifies outer retina changes in patients with biallelic RPE65 mutations undergoing gene therapy. Invest Ophthalmol Vis Sci 2025;66:5.

19. Eckardt F, Mittas R, Horlava N, et al. Deep Learning-Based Retinal Layer Segmentation in Optical Coherence Tomography Scans of Patients with Inherited Retinal Diseases. Klin Monbl Augenheilkd 2024. Available at: http://www.thieme-connect.de/DOI/DOI?10.1055/a-2227-3742 [Accessed June 25, 2025].

20. Woof WA, de Guimarães TAC, Al-Khuzaei S, et al. Quantification of fundus autofluorescence features in a molecularly characterized cohort of >3500 patients with inherited retinal disease from the United Kingdom. Ophthalmol Sci 2025;5:100652.

21. Mishra Z, Wang Z, Sadda SR, Hu Z. Using ensemble OCT-derived features beyond intensity features for enhanced Stargardt atrophy prediction with deep learning. Appl Sci 2023;13. Available at: 10.3390/app13148555.

22. Pontikos N, Arno G, Jurkute N, et al. Genetic Basis of Inherited Retinal Disease in a Molecularly Characterized Cohort of More Than 3000 Families from the United Kingdom. Ophthalmology 2020;127:1384–1394.

23. Lin S, Vermeirsch S, Pontikos N, et al. Spectrum of genetic variants in the commonest genes causing inherited retinal disease in a large molecularly characterised UK cohort. Ophthalmology Retina 2024. Available at: https://www.sciencedirect.com/science/article/pii/S2468653024000137.

24. Nguyen Q, Woof W, Kabiri N, et al. Can artificial intelligence accelerate the diagnosis of inherited retinal diseases? Protocol for a data-only retrospective cohort study (Eye2Gene). BMJ Open 2023;13:e071043.

25. Dice LR. Measures of the Amount of Ecologic Association Between Species. Ecology 1945;26:297–302.

26. Chollet F. Xception: Deep learning with depthwise separable convolutions. arXiv [csCV] 2016. Available at: http://arxiv.org/abs/1610.02357.

27. Isensee F, Jaeger PF, Kohl SAA, et al. nnU-Net: a self-configuring method for deep learning-based biomedical image segmentation. Nat Methods 2021;18:203–211.

28. Zhang G, Fu DJ, Liefers B, et al. Clinically relevant deep learning for detection and quantification of geographic atrophy from optical coherence tomography: a model development and external validation study. Lancet Digit Health 2021;3:e665–e675.

29. Hufnagel RB, Liang W, Duncan JL, et al. Tissue-specific genotype-phenotype correlations among USH2A-related disorders in the RUSH2A study. Hum Mutat 2022;43:613–624. Available at: [Accessed May 30, 2025].

30. Fujinami K, Lois N, Davidson AE, et al. A longitudinal study of stargardt disease: clinical and electrophysiologic assessment, progression, and genotype correlations. Am J Ophthalmol 2013;155:1075–1088.e13.

31. Fujinami K, Lois N, Mukherjee R, et al. A longitudinal study of Stargardt disease: quantitative assessment of fundus autofluorescence, progression, and genotype correlations. Invest Ophthalmol Vis Sci 2013;54:8181–8190.

32. Fujinami K, Zernant J, Chana RK, et al. Clinical and molecular characteristics of childhood-onset Stargardt disease. Ophthalmology 2015;122:326–334.

33. Fakin A, Robson AG, Fujinami K, et al. Phenotype and Progression of Retinal Degeneration Associated With Nullizigosity of ABCA4. Invest Ophthalmol Vis Sci 2016;57:4668–4678.

34. Tanna P, Georgiou M, Strauss RW, et al. Cross-Sectional and Longitudinal Assessment of the Ellipsoid Zone in Childhood-Onset Stargardt Disease. Transl Vis Sci Technol 2019;8:1.

35. Cornelis SS, Bauwens M, Haer-Wigman L, et al. Compendium of clinical variant classification for 2,247 unique ABCA4 variants to improve genetic medicine access for Stargardt Disease. 2023. Available at: 10.1101/2023.04.24.23288782.

36. Cornelis SS, Runhart EH, Bauwens M, et al. Personalized genetic counseling for Stargardt disease: Offspring risk estimates based on variant severity. Am J Hum Genet 2022;109:498– 507.

37. Lee W, Zernant J, Su P-Y, et al. A genotype-phenotype correlation matrix for ABCA4 disease based on long-term prognostic outcomes. JCI Insight 2022;7. Available at: 10.1172/jci.insight.156154.

38. Fujinami K, Waheed N, Laich Y, et al. Stargardt macular dystrophy and therapeutic approaches. Br J Ophthalmol 2024;108:495–505.

39. Georgiou M, Kane T, Tanna P, et al. Prospective Cohort Study of Childhood-Onset Stargardt Disease: Fundus Autofluorescence Imaging, Progression, Comparison with Adult-Onset Disease, and Disease Symmetry. Am J Ophthalmol 2020;211:159–175.

40. Loo J, Jaffe GJ, Duncan JL, et al. Validation of a deep learning-based algorithm for segmentation of the ellipsoid zone on optical coherence tomography images of an ush2a-related retinal degeneration clinical trial. Retina 2022;42:1347–1355.

41. Wang Y-Z, Wu W, Birch DG. A Hybrid Model Composed of Two Convolutional Neural Networks (CNNs) for Automatic Retinal Layer Segmentation of OCT Images in Retinitis Pigmentosa (RP). Transl Vis Sci Technol 2021;10:9.

42. Khan KN, Robson A, Mahroo OAR, et al. A clinical and molecular characterisation of CRB1-associated maculopathy. Eur J Hum Genet 2018;26:687–694.

43. Daich Varela M, Georgiou M, Alswaiti Y, et al. CRB1-associated retinal dystrophies: Genetics, clinical characteristics, and natural history. Am J Ophthalmol 2023;246:107–121.

44. Laich Y, Georgiou M, Fujinami K, et al. Best vitelliform macular dystrophy natural history study report 1: Clinical features and genetic findings. Ophthalmology 2024;131:845–854.

45. de Guimarães TAC, Kalitzeos A, Mahroo OA, et al. A long-term retrospective natural history study of EFEMP1-associated autosomal dominant drusen. Invest Ophthalmol Vis Sci 2024;65:31.

46. De Silva SR, Arno G, Robson AG, et al. The X-linked retinopathies: Physiological insights, pathogenic mechanisms, phenotypic features and novel therapies. Prog Retin Eye Res 2021;82:100898.

47. Tee JJL, Smith AJ, Hardcastle AJ, Michaelides M. RPGR-associated retinopathy: clinical features, molecular genetics, animal models and therapeutic options. Br J Ophthalmol 2016;100:1022–1027.

